# Estimating the impact of vaccination and long-acting monoclonal antibodies for RSV epidemics across Hong Kong, Beijing, and Thailand: a modelling study

**DOI:** 10.1101/2025.11.06.25339729

**Authors:** Kathy Leung, Ivan F. N. Hung, Chelsea L. Hansen, Carolyn A. Cohen, Jiayi Du, Hui-Ling Yen, Sophie A. Valkenburg, J. S. Malik Peiris, Cecile Viboud, Joseph T. Wu

## Abstract

**Background:** Respiratory syncytial virus (RSV) is a major cause of acute lower respiratory infections worldwide, with high morbidity among infants and older adults. Recent approvals of long-acting monoclonal antibodies (mAbs) and vaccines offer new prevention opportunities, but their impact in Asian settings remains uncertain.

**Methods:** We developed age-structured SEIR transmission models for Hong Kong, Beijing, and Thailand, calibrated to region-specific surveillance and seroprevalence data from 2014–2019. Using Bayesian inference, we estimated RSV transmission dynamics and simulated intervention scenarios involving long-acting mAbs for high-risk infants, maternal vaccination, and older adult vaccination.

**Findings:** RSV showed distinct seasonality: year-round in Hong Kong, winter peaks in Beijing, and rainy-season peaks in Thailand. Estimated annual infection attack rates among infants <1 year were 51.1% in Hong Kong and 22.5% in Beijing, and 75.8% and 70.1% among children aged 1–4 years, compared with 27.9% among 0–4 years in Thailand. Simulations suggest long-acting mAbs and maternal vaccination (coverage 38.5%) could avert 15.6–19.5% and 18.0–25.2% of severe infant outcomes, respectively. Vaccination of older adults (coverage 30–40%) reduced RSV-associated outcomes by 21.7–27.7% in Hong Kong, 33.9–39.7% in Beijing and 34.8–49.8% in Thailand. Combined interventions achieved reductions of 39.2% (27.5–55.8), 49.4% (42.0–59.7), and 53.8% (44.1–64.8) in severe outcomes among infants <1 year, 19.3% (18.9–21.7), 27.7% (26.9–31.5) and 31.5% (31.3–33.9) among 60-74 years, 26.5% (26.2–28.7), 52.3% (51.5–54.8) and 37.8% (37.6–40.0) among ≥75 years, in Hong Kong, Beijing, and Thailand, respectively.

**Interpretation:** Our modelling framework provides a novel approach to evaluate RSV prevention strategies in Asian populations with diverse seasonality. As real-world effectiveness data emerge, future research should refine estimates and optimise intervention combinations for maximum public health impact.

**Funding:** MSD Investigator Studies Program, General Research Fund, Health and Medical Research Fund. (Word count: 298)

**Research in context:** *Evidence before this study:* We searched PubMed and Google Scholar for studies published before October 2025 using terms such as *“RSV transmission model,” “RSV monoclonal antibodies,” “RSV maternal vaccination,”* and *“older adult RSV vaccine.”* Most modelling studies focused on high-income countries in temperate regions, including Europe, the United States, and Australia. These studies evaluated the impact of RSV immunization strategies but did not address populations with diverse seasonality patterns or those in low- and middle-income countries. Evidence on RSV burden in Asia remains limited, and few studies have integrated seroprevalence data with transmission modelling to inform intervention strategies.

*Added value of this study:* This study provides the first comparative modelling analysis of RSV transmission dynamics and intervention impacts across three Asian settings—Hong Kong, Beijing, and Thailand— representing subtropical, temperate, and tropical climates. By calibrating age-structured SEIR models to local surveillance and seroprevalence data, we estimated infection attack rates and assessed the potential benefits of long-acting monoclonal antibodies, maternal vaccination, and older adult vaccination. Our findings show that moderate uptake of RSV interventions can substantially reduce severe outcomes and reinforce the need for implementing comprehensive prevention strategies in Asian settings.

*Implications of all the available evidence:* Our modelling framework offers a practical tool for policymakers to evaluate intervention combinations and prioritise resource allocation. As real-world effectiveness and uptake data become available, these insights can guide optimisation of RSV prevention programmes across diverse settings.

## Introduction

Respiratory syncytial virus (RSV) is a major global cause of hospitalizations and deaths across age groups, particularly impacting infants, young children, and older adults [1]. In 2019, worldwide estimates attributed 33.0 million RSV-associated acute lower respiratory infection (ALRI) episodes, 3.6 million ALRI hospitalizations, and 101,400 RSV-attributable deaths to children aged 0-5 years [2]. RSV also imposes a substantial disease burden on older adults, particularly those with comorbidities and underlying risk factors [3]. While robust surveillance data on RSV epidemiology and the associated health impacts are available in many high-income countries, including European nations [4, 5], Australia [6], and the United States [7], most low- and middle-income countries (LMICs) in Africa and Asia possess limited or no national RSV surveillance data [8, 9], though RSV disease burden is substantial in these countries, such as China and Thailand [2, 3, 10].

In mainland China, a 2015 systematic review reported RSV in 18.7% of acute respiratory tract infections (ARI), with prevalence ranging from 2.8% in adolescents and adults to 26.5% among infants aged below 6 months [11]. Hong Kong has also observed a significant RSV burden, with the annual cumulative hospitalization rate for all ages ranging from 45 to 97 per 100,000 population between 2016 and 2024 [12]. It was estimated that RSV accounted for 37% of respiratory viral infection hospitalizations in children under five [13], and reported an annual RSV hospitalization rate of 233.4 to 311.2 per 10,000 in infants <6 months. In Thailand, 8.1% of hospitalized ALRI patients tested RSV-positive between 2008 and 2011, with an estimated incidence of 154.3 RSV-associated ALRI hospitalizations per 10,000 person-years among children under 1 year and 98.1 per 10,000 among those under 5 years [14]. Despite limited testing capacity, national data from 2015 to 2020 indicate that RSV-related lower respiratory tract infections (LRTIs) accounted for at least 1.2% (19,340 of 1,610,160) of all hospital admissions in children under two years of age, with RSV pneumonia causing over 70% of these admissions [15].

Efforts to develop vaccines and immunoprophylaxis for preventing paediatric RSV infections have been highly active in recent years [16]. Prior to 2023, no RSV prophylaxis was available for adults, and palivizumab was the sole prophylaxis against severe RSV disease in infants, reserved exclusively for very premature newborns and those with high-risk conditions [17]. However, the landscape of RSV prevention changed substantially in 2023–2025 with the approvals of nirsevimab and clesrovimab, both long-acting monoclonal antibodies (mAb) for routine infant use, which provides protection lasting at least 5–6 months [18]. In parallel, three vaccines for high-risk and older adults were approved, including one also recommended for use in pregnant women to protect newborns [19].

With the expanded options and availability of RSV vaccines and mAbs, it is essential to characterize RSV epidemiology and estimate the associated disease burden to inform optimal vaccination and prophylaxis strategies. In this study, we leveraged published and routinely collected surveillance data to develop an age-specific RSV transmission model for three Asian populations: Hong Kong, Beijing, and Thailand. We used this model to estimate the number of RSV-related ALRIs and/or hospitalizations averted among infants and older adults aged 60 or above. Given the current limited availability of RSV vaccines and mAbs in these regions, we assumed uptake and effectiveness parameters based on real-world evidence from the United States, applied prospectively to simulate post-introduction impact [20]. The model incorporated region-specific demographic structures, age-dependent contact patterns, and locally calibrated disease burden estimates to compare the potential public health impact of alternative vaccination and mAb prophylaxis strategies across the three settings.

## Methods

### Age-stratified hospital admission or laboratory-confirmed case data

Given that the transmission of many respiratory diseases was suppressed by public health and social measures implemented during the COVID-19 pandemic [21, 22], our analysis first focused on weekly or monthly data during the pre-pandemic period from 2014 to 2019 to estimate the RSV transmission dynamics. For Hong Kong, we characterized RSV epidemiology using weekly counts of laboratory-confirmed RSV cases with monthly age stratifications reported by the Centre for Health Protection between June 2015 and December 2019 [12]. Data from February 2020 to December 2024 were subsequently used to estimate the impacts of public health and social measures (PHSMs) on RSV transmission in Hong Kong [22]. For Beijing, monthly RSV case numbers were obtained from the Respiratory Pathogen Surveillance System (RPSS) sentinel network, which comprises 35 sentinel hospitals; age-stratified case numbers were only available annually [23]. Similarly, for Thailand, we obtained monthly numbers of RSV-related LRTI hospital admissions in children under 18 from Sitthikarnkha *et al* [15]. Given the lack of national adult data in Thailand, we estimated RSV-related LRTI hospital admissions among individuals aged 18 or above by applying age-group specific ratios of RSV-associated ALRI hospitalizations from Naorat *et al* [14].

### Seroprevalence data

Serological data on RSV in Asian populations remain limited, with published studies available only for Beijing and Thailand. We incorporated cross-sectional seroprevalence data from Beijing collected in 2008 by Lu *et al*., comprising 278 children aged 1 month to 5 years, 122 individuals aged 6–19 years, and 756 adults aged 20–60 years [24]. In Thailand, Pasittungkul *et al*. collected longitudinal samples from 302 mothers and 291 newborns between 2015 and 2021, with specimens obtained at 2, 7, 18, 19, 24, 36, 48, and 60 months of age [25]. To fill the data gap for Hong Kong, we conducted RSV seroprevalence testing with ELISA on 299 serum samples collected between April 2020 and March 2021 from individuals aged 3 months to 71 years [26]. We further supplemented our analysis with aggregate seroprevalence estimates, specifically from neutralization assays or IgG ELISA, derived from the meta-analysis by Nakajo and Nishiura [27]. Although RSV epidemiology exhibits geographic and temporal heterogeneity with distinct seasonality [28], most seroprevalence studies demonstrate similar patterns: seroprevalence is lowest among infants aged 0–1 year, increases during early childhood, and reaches a high plateau in older children and adults [27]. We therefore integrated all available seroprevalence data into the calibration of each region-specific transmission model.

### RSV transmission model

We adapted the age-structured deterministic SEIR (susceptible-exposed-infectious-removed) compartmental model for RSV transmission, originally developed by Hodgson *et al* [4]. This model incorporates key RSV immunological features: short-term maternally derived immunity, lifelong repeated infections, a brief period of full post-infection protection, and the gradual acquisition of partial permanent immunity (**Figure S1**). The model allows parameterization for individual regions using their specific demographic and age-dependent contact data [29–31]. A detailed model diagram and all parameter descriptions are provided in the **Supplementary Information**.

We used a Bayesian Markov chain Monte Carlo (MCMC) approach to fit each region-specific transmission model to the previously described age-stratified confirmed case or hospital admission data from each region but all available seroprevalence data. While demographic and contact pattern parameters were fixed based on published literature, we estimated local-specific susceptibility, infectiousness, seasonality, immunity, and reporting rates by fitting the model to our data (**Supplementary Information**).

### RSV interventions with mAbs and vaccinations

*Baseline scenario.* Prior to 2022, none of the three regions had established a local consensus on the use of mAbs for RSV prophylaxis [32]. Although Hong Kong experts reached agreement in July 2022 to consider a 6-month regimen of mAb prophylaxis for preterm infants (<29 weeks’ gestational age) and high-risk infants under the age of 1 year [33], the coverage of mAb was minimal in Hong Kong before 2022, and no formal regional recommendations for RSV prophylaxis using mAbs or vaccines had been adopted in any of the three settings by the end of 2024. Accordingly, in this study, we assumed no administration of RSV mAbs or vaccines across any age group in any region during the baseline period (2014–2019). This baseline scenario serves as the reference point against which we evaluate the projected public health impact of alternative intervention strategies (**Figure S2**).

*Long acting mAb use.* Based on an observational study of over 9 million Chinese women (2012-2018), we assumed that 6.4% of infants born preterm would receive long acting mAb (e.g., nirsevimab, which lasts 5-6 months with one dose) [34]. Following estimates by Wang *et al*., we assumed that preterm infants accounted for 25% (95% uncertainty range: 16–37) of severe outcomes (e.g., hospitalizations) due to RSV-associated acute lower respiratory infection (ALRI) among all infants, regardless of gestational age [35]. Based on results from the MELODY trial, we assumed the long-acting mAb provided 77.3% efficacy (95% CI: 50.3 – 89.7) against RSV-associated hospitalizations for a duration of 180 days post-administration (see **Supplementary Information**) [36].

*Maternal vaccination.* We assumed that 38.5% (95% CI: 23.2 – 53.5) of pregnant women would receive maternal vaccination, based on early 2025 data from the U.S. CDC [37]. For these vaccinated mothers, we parameterised the model using Abrysvo’s efficacy against severe lower respiratory tract infection (LRTI), estimated at 69.4% (95% CI: 44.3–84.1) for an average duration of 180 days after birth (see **Supplementary Information**) [38]. When both long-acting mAbs and maternal vaccination were considered, we assumed that preterm infants would receive either long-acting mAbs or be covered by maternal vaccination through their mothers.

*Vaccinating older adults.* As subsidized RSV vaccination is currently unavailable for older adults in the three regions, we assumed vaccination coverage of 39.7% (95% CI: 23.1 – 58.9) among individuals aged ≥75 years and 31.4% (95% CI: 18.5 – 45.7) among those aged 60– 74 years, based on US CDC data [39]. For reference, seasonal influenza vaccine uptake in Hong Kong during 2024–2025 was 25.6% among individuals aged 50–64 years and 51.8% among those aged ≥65 years [40], compared with 48.4% and 71.8%, respectively, in the United States [41]. In Thailand, influenza vaccine coverage among older adults was approximately 47% in 2024, whereas a recent study reported substantially lower influenza vaccine uptake among older adults in Beijing: 63% sporadically vaccinated, 19% occasionally vaccinated, and 18% frequently vaccinated [42, 43]. We parameterized the model for protection against adult hospitalizations using real-world vaccine effectiveness estimates of 80% (95% CI: 71–85) from the U.S. during the 2023–2024 season [44], and assumed the same efficacy for protection against infection (see **Supplementary Information**).

## Results

### RSV epidemiology in Hong Kong, Beijing and Thailand

Our model reproduced the age distribution and seasonality patterns of RSV hospital admissions and laboratory-confirmed cases across all three regions (Hong Kong, Beijing, and Thailand) between 2015 and 2019 (**Figures 1-4, Figures S3-S5**). By calibrating the model to both RSV case data and seroprevalence data, we were able to estimate transmission dynamics parameters not directly available from observational studies (**Tables S1-S4**). In Hong Kong, RSV circulated year-round, exhibiting an irregular seasonal pattern with a larger peak typically in September/October and a smaller peak in February/March in some years (**Figure 1-2**). In contrast, RSV in Beijing followed a typical winter seasonal pattern, consistent with most of Northern China [45], with the peak usually occurring in December/January (**Figure 3**). In Thailand, RSV circulated year-round, but the RSV season coincided with the rainy season, peaking in August or September (**Figure 4**).

**Figure 1.**
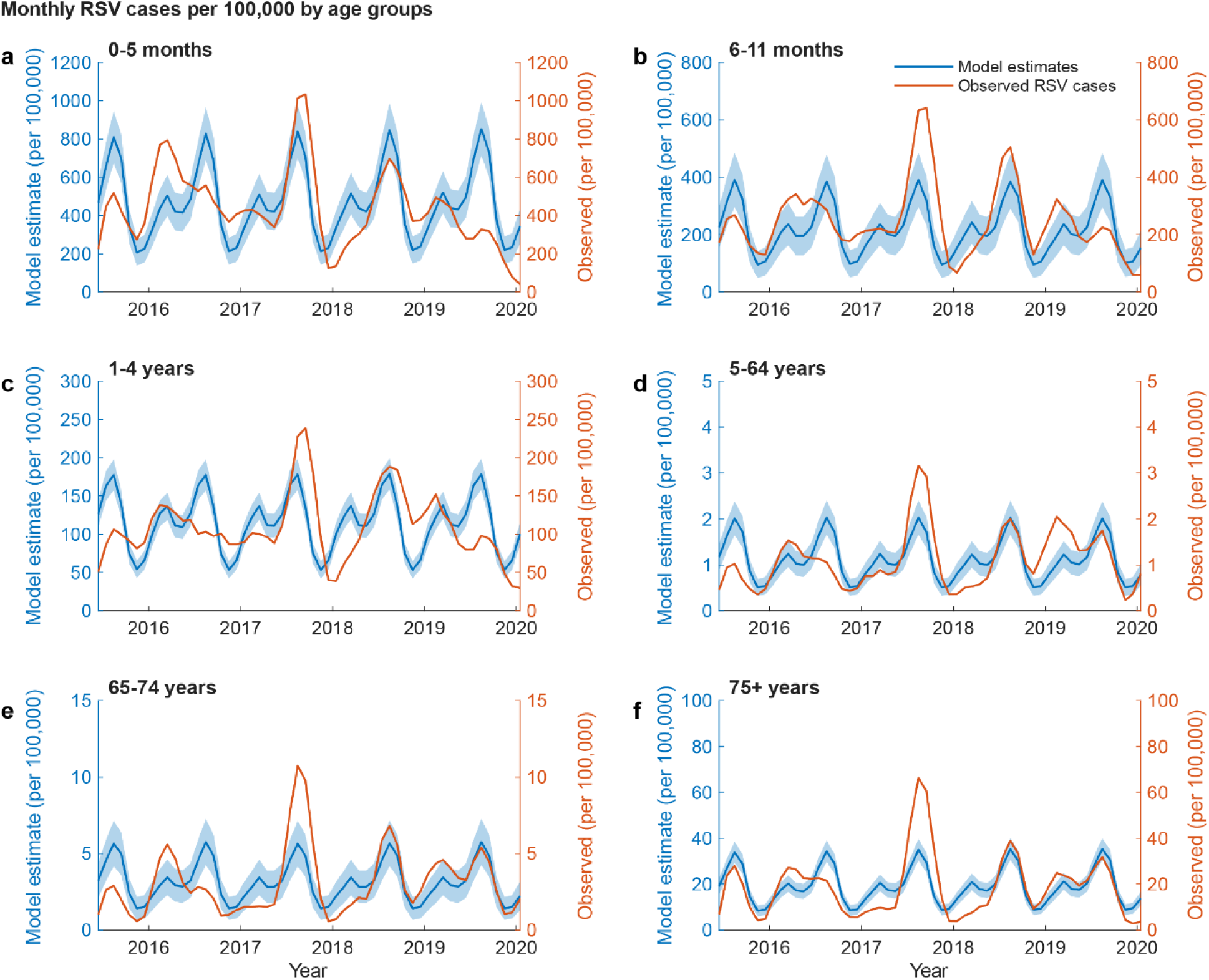
Monthly number of laboratory-confirmed RSV cases per 100,000 population by age in Hong Kong between 1 June 2015 to 22 January 2020. (a) 0-5 months. (b) 6-11 months. (c) 1-4 years. (d) 5-64 years. (e) 65-74 years. (f) ≥ 75 years. Blue solid lines indicated posterior mean, and blue shades indicated 95% CrI. Orange lines indicated the observed monthly number of laboratory-confirmed RSV cases per 100,000 population (3-month moving average accounting for reporting delays [48]). The x-axis labels indicated January 1 of each year.

**Figure 2.**
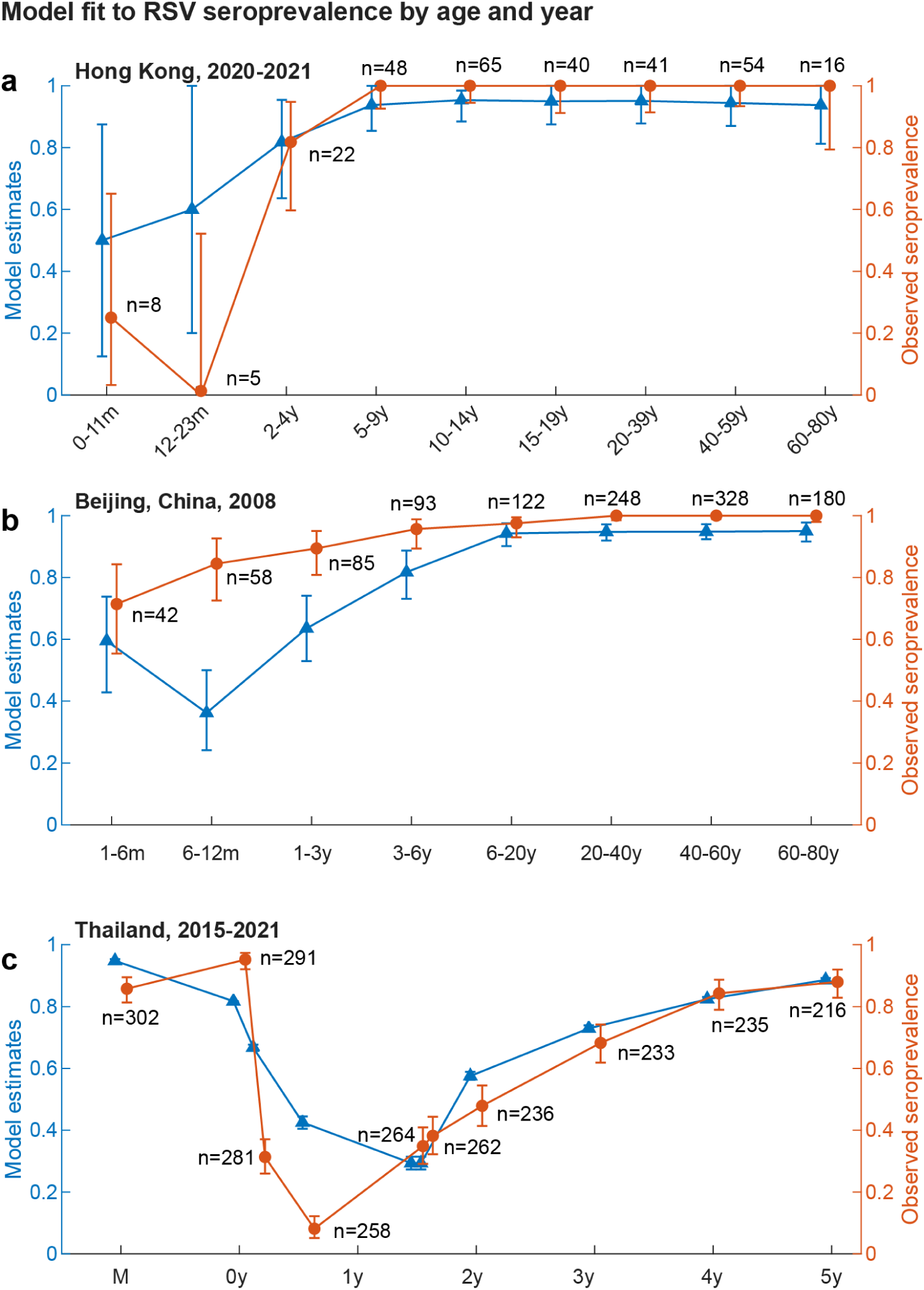
Estimated RSV seroprevalence by age in Hong Kong between 2014 and 2019 and model fit to seroprevalence data in Beijing, Hong Kong and Thailand. The age-specific seroprevalences from the Hong Kong transmission model were fit to all available seroprevalence data [17–19] (See Supplementary Information). Model fit was shown to (a) seroprevalence from Hong Kong, with samples collected in 2020-2021 from individuals aged 3 months to 71 years; (b) seroprevalence from Beijing, with samples collected in 2008 from individuals aged 1-6 months to 60-80 years; (c) seroprevalence from Thailand, with samples collected between 2015 and 2021 from mothers (indicated by “M”), newborn infants, and children aged 2 months to 5 years. Blue triangles indicated the posterior mean, and blue bars indicated 95% CrI. Orange dots indicated the observed seroprevalence, and orange bars indicated the 95% CI assuming the number of seropositive samples follow binomial distribution. The number next to each orange dot indicated the sample size of the age group of interest in the data.

**Figure 3.**
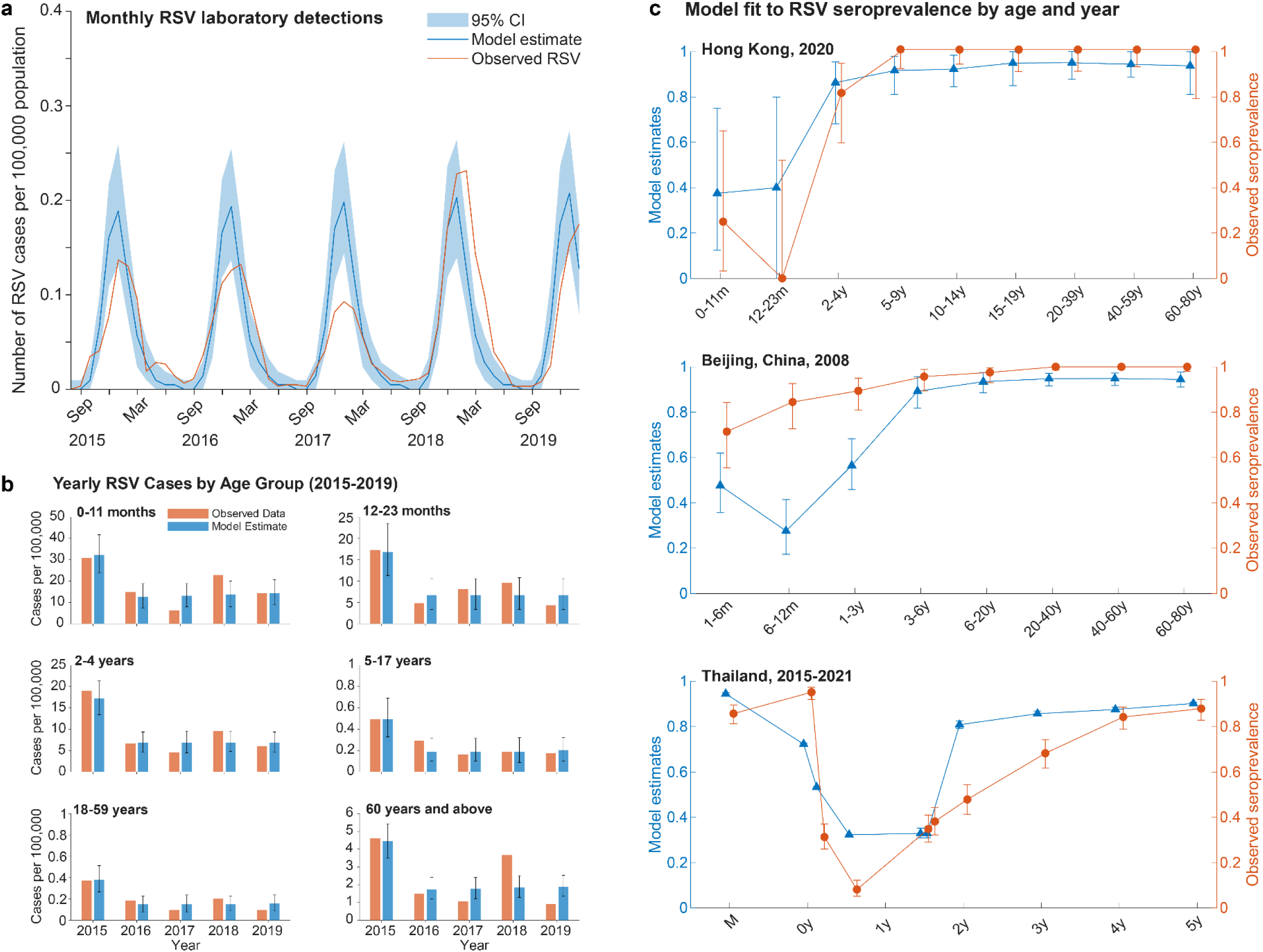
RSV-associated acute respiratory tract infections per 100,000 population in Beijing reported by the RPSS sentinel network between July 2015 to December 2019. (a) Monthly number of laboratory-confirmed RSV-associated acute respiratory tract infections per 100,000 population. Blue solid lines indicated the posterior mean, and blue shades indicated 95% CrI. Orange lines indicated the observed monthly number of RSV-associated acute respiratory tract infections per 100,000 population (3-month moving average accounting for reporting delays [23]). (b) Yearly number of RSV-associated acute respiratory tract infections by age reported between 2015 and 2019. Blue bars indicated the posterior mean with 95% CrI and the orange bars indicated the observed number of RSV-associated acute respiratory tract infections [23]. (c) Seroprevalence data from Hong Kong, Beijing and Thailand. Blue triangles indicated the posterior mean, and blue bars indicated 95% CrI. Orange dots indicated the observed seroprevalence, and orange bars indicated the 95% CI assuming the number of seropositive samples follow binomial distribution.

**Figure 4.**
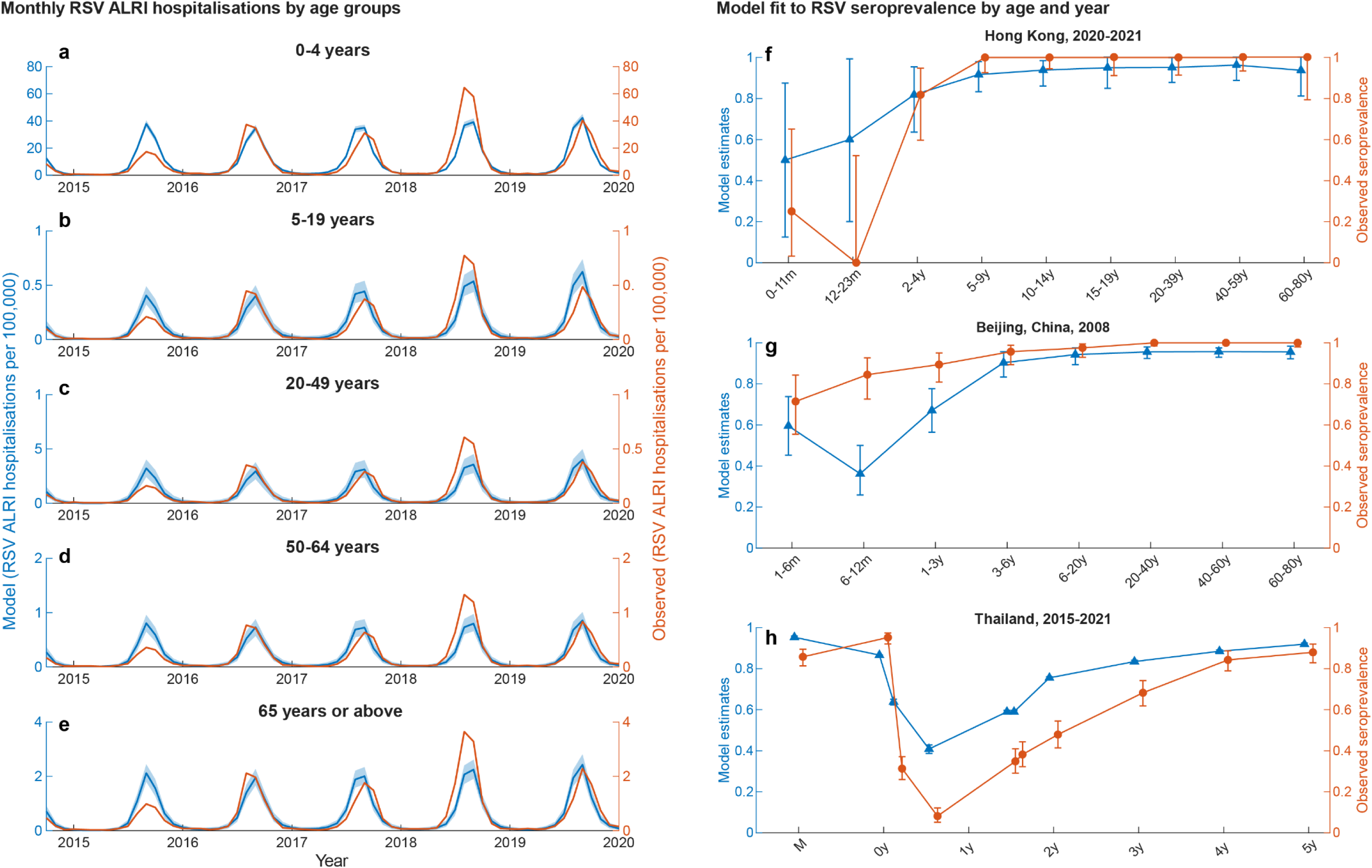
Monthly number of hospitalisations for RSV-associated acute lower respiratory tract infections per 100,000 population by age between October 2014 and December 2019 and estimated age-specific seroprevalence in Thailand. Monthly number of hospitalisations for RSV-associated ALRI hospitalisation of individuals aged (a) 0-4 years, (b) 5-19 years, (c) 20-49 years, (d) 50-64 years, (e) ≥ 65 years. Blue solid lines indicated posterior mean, and blue shades indicated 95% CrI. Orange lines indicated the observed monthly number of hospitalisations for RSV-associated acute lower respiratory tract infections per 100,000 population. The x-axis labels indicated January 1 of each year. Model fit was shown to seroprevalence from (f) Hong Kong, (g) Beijing, and (h) Thailand. Blue triangles indicated the posterior mean, and blue bars indicated 95% CrI. Orange dots indicated the observed seroprevalence, and orange bars indicated the 95% CI assuming the number of seropositive samples follow binomial distribution.

Our model indicated that several transmission dynamics parameters, including immunity duration and relative susceptibility after the first infection, were associated with the regional RSV seasonality (**Table S3**). For example, the estimated susceptibility to secondary infection relative to primary infection was 0.91 in Hong Kong—substantially higher than in Beijing (0.20) and Thailand (0.05)—corresponding to Hong Kong’s year-round RSV circulation and a higher infection attack rate among infants younger than 1 year (**Table 1**). Consequently, by age two, children in Hong Kong were typically infected twice. The average ages of the first, second, and third infections were 1.39 (1.27–1.62), 2.20 (2.10–2.41), and 2.95 (2.86–3.16) in Hong Kong; 2.14 (2.09–2.48), 2.76 (2.66–3.15), and 3.38 (3.32–3.63) in Beijing; and 1.95 (1.89–1.99), 2.57 (2.54–2.64), and 3.33 (3.31–3.40) in Thailand, respectively. Across the three regions, the estimated mean post-infection immunity duration ranged from 117 to 216 days (**Table S3**), consistent with the 2–6 months reported in experimental challenge studies [46]. In contrast, the estimated duration of maternally derived immunity was much longer (243–295 days), likely reflecting its dependence on both regional RSV seasonality and the prevalence of recently infected mothers at birth.

**Table 1.**
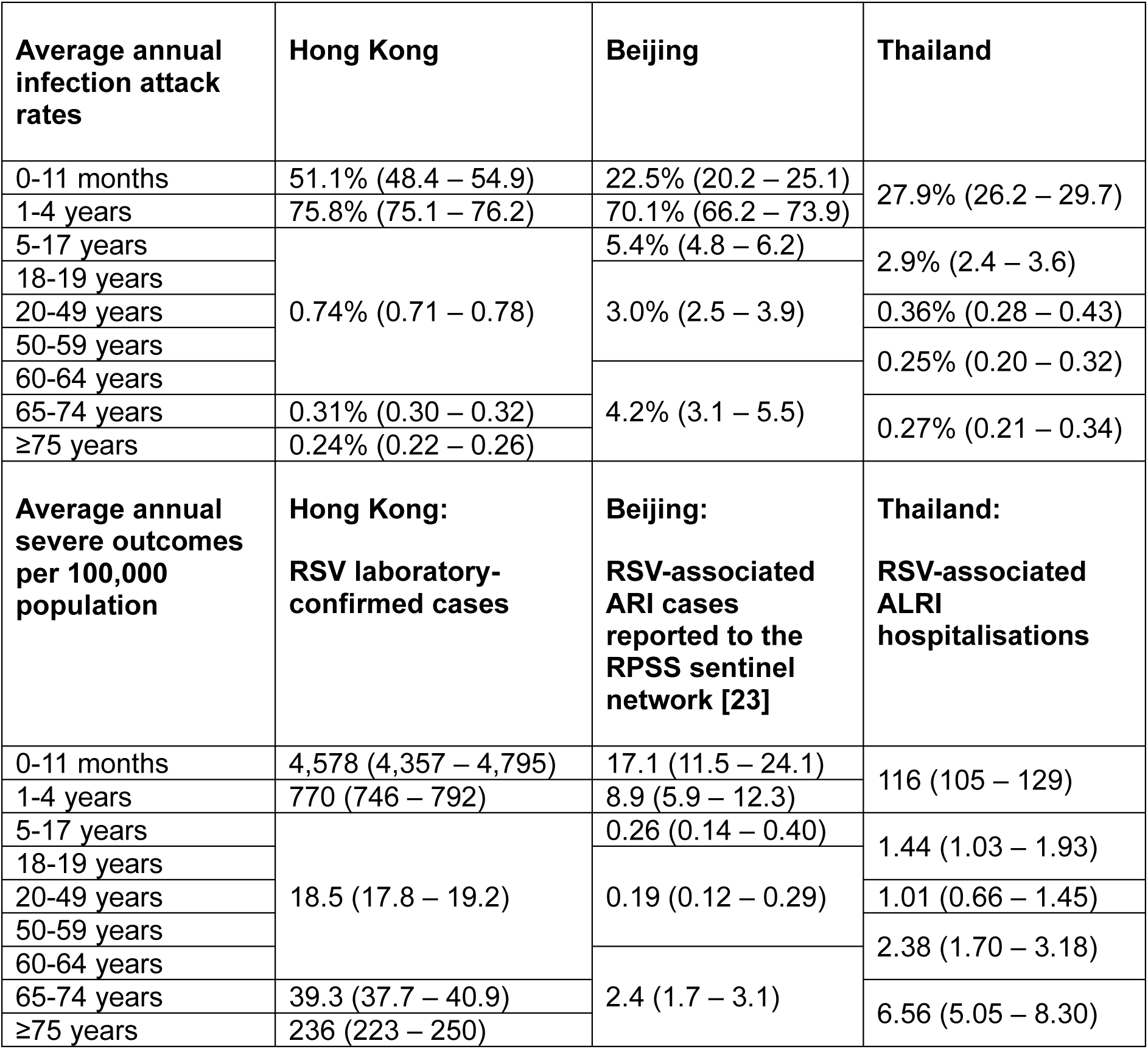
Estimated RSV burden in Hong Kong, Beijing and Thailand.

### RSV disease burden

In Hong Kong and Beijing, we estimated that 51.1% (95% CrI: 48.4–54.9) and 22.5% (95% CrI: 20.2–25.1) of children, respectively, were infected with RSV during their first year of life (**Table 1**). Among those infected, infants under 1 year had the highest probability of being laboratory-confirmed or reported to RPSS. Annual infection attack rates remained high among children under five, ranging from 27.9% (95% CrI: 26.2–29.7) in Thailand to 75.8% (95% CrI: 75.1–76.2) and 70.1% (95% CrI: 66.2–73.9) among children aged 1–4 years in Hong Kong and Beijing, respectively. The annual attack rates declined progressively with age among adolescents, adults, and older adults, with the lowest rates observed in individuals aged ≥65 years: 0.24–0.31% in Hong Kong, 4.2% in Beijing, and 0.27% in Thailand (**Table 1**).

The average probability of an RSV infection being laboratory-confirmed was highest in Hong Kong among children younger than six months and adults aged 75 years or older (**Table S3**). The annual number of RSV laboratory-confirmed cases per 100,000 population in Hong Kong was 4,578 (95% CrI: 4,357-4,795) for individuals aged 0-11 months, 770 (95% CrI: 746-792) for those 1-4 years, 18.5 (95% CrI: 17.8-19.2) for 5-64 years, 39.3 (95% CrI: 37.7-40.9) for 65-74 years, and 236 (95% CrI: 223-250) for individuals 75 years or above (**Table 1**).

Similarly, the probability of RSV infection being reported to RPSS in Beijing and the probability of developing RSV-associated ALRI hospitalizations in Thailand were highest among children under six months and the elderly aged 60 or above (**Table 1**). The annual number of RSV-associated ALRI hospitalizations per 100,000 population in Thailand was 116 (95% CrI: 105-129) for individuals aged 0-4 years, 1.44 (95% CrI: 1.03-1.95) for 5-19 years, 1.01 (95% CrI: 0.66-1.45) for 20-49 years, 2.38 (95% CrI: 1.70-3.18) for 50-64 years, and 6.56 (95% CrI: 5.05-8.30) for individuals 75 years or above.

Due to public health and social measures (PHSMs) implemented in response to the COVID-19 pandemic [47], we estimated a substantial reduction in RSV activity in Hong Kong between 2020 and 2024 (**Figure S6**). Following the declaration of the pandemic on January 23, 2020, RSV activity declined sharply across all age groups through February 28, 2022, with reductions of 98.0% (96.8–98.9%), 80.5% (79.3–81.7%), 98.8% (97.5–99.8%), 99.7% (98.7–100%), and 99.9% (99.6–100%) among individuals aged 0–11 months, 1–4 years, 5–64 years, 65–74 years, and ≥75 years, respectively. During the major fifth wave of the pandemic (March 1 to August 31, 2022), RSV circulation remained minimal (**Table S4**). RSV activity subsequently showed a modest increase. Between September 1, 2022, and May 31, 2024, compared to the pre-pandemic period, RSV activity remained markedly reduced among children aged 0–11 months and 1–4 years, with declines ranging from 78.3% to 97.1%. However, a rebound in RSV transmission was observed among adults following the full relaxation of PHSMs. In the summer and autumn of 2023, estimated RSV transmission among adults aged 5–64 and 65–74 years increased substantially compared with pre-pandemic levels. Similarly, in spring 2024, infection rates among individuals aged ≥65 years consistently exceeded pre-pandemic levels (**Figure S6**).

### Impact of RSV interventions with mAbs and vaccinations

We applied the calibrated region-specific models to simulate RSV interventions using long-acting mAbs and vaccinations targeting infants, pregnant women, and older adults (**Table 2**). Compared to a baseline scenario without interventions, administering long-acting mAbs to high-risk infants and maternal vaccination (with an estimated uptake of ∼38.5%) achieved similar reductions in RSV-associated cases, ARIs, and hospitalizations (**Table 2**). For example, in Hong Kong, the annual number of averted RSV laboratory-confirmed cases among children under 1 year was 667 (95% CrI: 427–1,015) for mAbs and 765 (95% CrI: 400–1,155) for maternal vaccination. Increasing maternal vaccination coverage to 75% and 95% in Hong Kong further reduced severe outcomes among infants from 18.0% to 29.3% and 44.0%, respectively. In Thailand, the estimated number of averted RSV-associated ALRI hospitalizations was 93 (95% CrI: 71–128) for mAbs and 120 (95% CrI: 90–152) for maternal vaccination. Across the three regions, long-acting mAbs reduced severe outcomes among infants younger than 1 year by 15.6–19.5%, while maternal vaccination reduced severe outcomes by 18.0–25.2%.

**Table 2.**
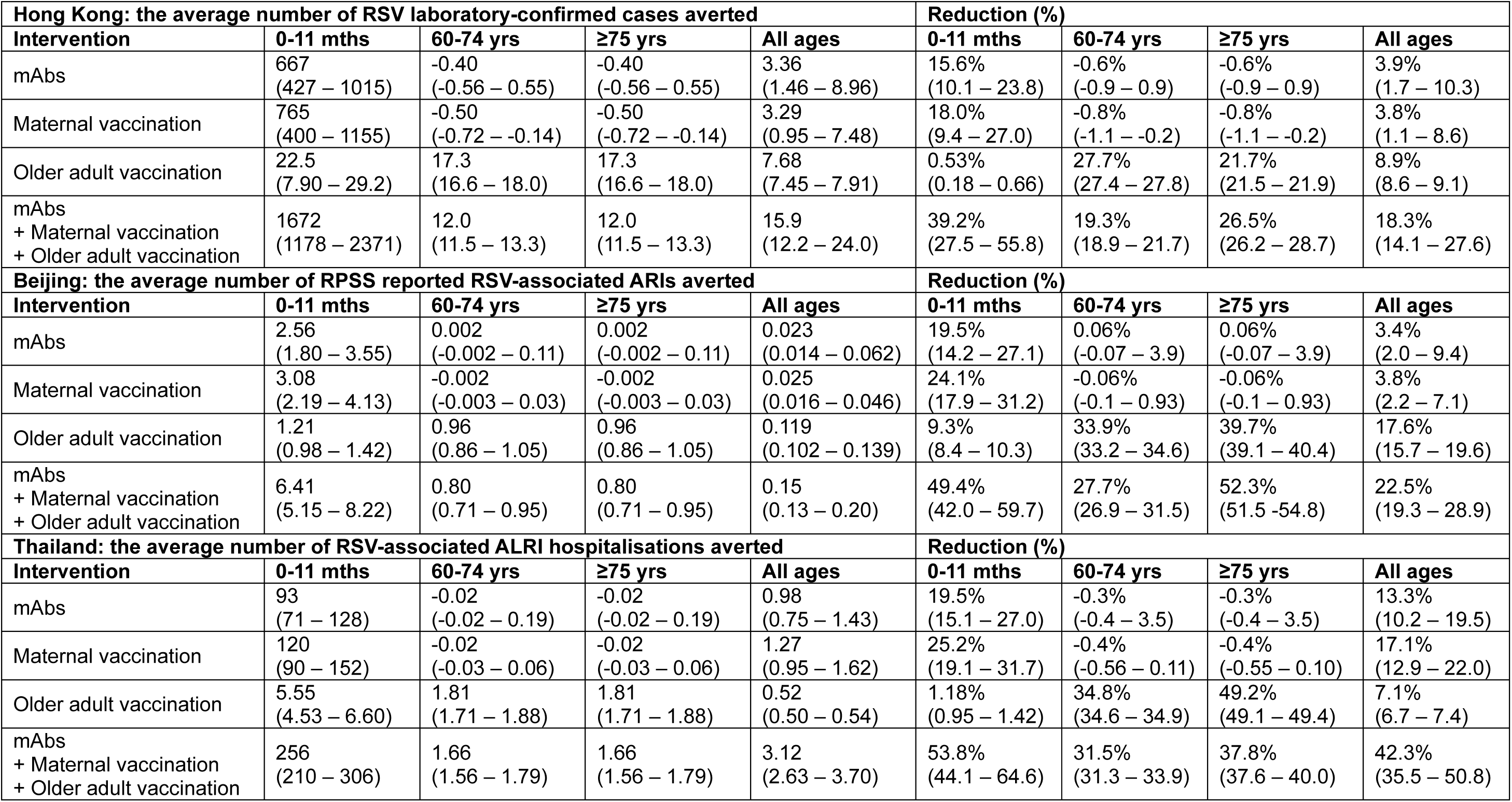
The impacts of RSV mAb administration and vaccinations in Hong Kong, Beijing and Thailand per 100,000 population.

| **Hong Kong: the average number of RSV laboratory-confirmed cases averted** |  |  |  |  | **Reduction (%)** |  |  |  |
| --- | --- | --- | --- | --- | --- | --- | --- | --- |
| **Intervention** | **0-11 mths** | **60-74 yrs** | **≥75 yrs** | **All ages** | **0-11 mths** | **60-74 yrs** | **≥75 yrs** | **All ages** |
| mAbs | 667<br>(427 – 1015) | -0.40<br>(-0.56 – 0.55) | -0.40<br>(-0.56 – 0.55) | 3.36<br>(1.46 – 8.96) | 15.6%<br>(10.1 – 23.8) | -0.6%<br>(-0.9 – 0.9) | -0.6%<br>(-0.9 – 0.9) | 3.9%<br>(1.7 – 10.3) |
| Maternal vaccination | 765<br>(400 – 1155) | -0.50<br>(-0.72 – -0.14) | -0.50<br>(-0.72 – -0.14) | 3.29<br>(0.95 – 7.48) | 18.0%<br>(9.4 – 27.0) | -0.8%<br>(-1.1 – -0.2) | -0.8%<br>(-1.1 – -0.2) | 3.8%<br>(1.1 – 8.6) |
| Older adult vaccination | 22.5<br>(7.90 – 29.2) | 17.3<br>(16.6 – 18.0) | 17.3<br>(16.6 – 18.0) | 7.68<br>(7.45 – 7.91) | 0.53%<br>(0.18 – 0.66) | 27.7%<br>(27.4 – 27.8) | 21.7%<br>(21.5 – 21.9) | 8.9%<br>(8.6 – 9.1) |
| mAbs<br>+ Maternal vaccination<br>+ Older adult vaccination | 1672<br>(1178 – 2371) | 12.0<br>(11.5 – 13.3) | 12.0<br>(11.5 – 13.3) | 15.9<br>(12.2 – 24.0) | 39.2%<br>(27.5 – 55.8) | 19.3%<br>(18.9 – 21.7) | 26.5%<br>(26.2 – 28.7) | 18.3%<br>(14.1 – 27.6) |
| **Beijing: the average number of RPSS reported RSV-associated ARIs averted** |  |  |  |  | **Reduction (%)** |  |  |  |
| **Intervention** | **0-11 mths** | **60-74 yrs** | **≥75 yrs** | **All ages** | **0-11 mths** | **60-74 yrs** | **≥75 yrs** | **All ages** |
| mAbs | 2.56<br>(1.80 – 3.55) | 0.002<br>(-0.002 – 0.11) | 0.002<br>(-0.002 – 0.11) | 0.023<br>(0.014 – 0.062) | 19.5%<br>(14.2 – 27.1) | 0.06%<br>(-0.07 – 3.9) | 0.06%<br>(-0.07 – 3.9) | 3.4%<br>(2.0 – 9.4) |
| Maternal vaccination | 3.08<br>(2.19 – 4.13) | -0.002<br>(-0.003 – 0.03) | -0.002<br>(-0.003 – 0.03) | 0.025<br>(0.016 – 0.046) | 24.1%<br>(17.9 – 31.2) | -0.06%<br>(-0.1 – 0.93) | -0.06%<br>(-0.1 – 0.93) | 3.8%<br>(2.2 – 7.1) |
| Older adult vaccination | 1.21<br>(0.98 – 1.42) | 0.96<br>(0.86 – 1.05) | 0.96<br>(0.86 – 1.05) | 0.119<br>(0.102 – 0.139) | 9.3%<br>(8.4 – 10.3) | 33.9%<br>(33.2 – 34.6) | 39.7%<br>(39.1 – 40.4) | 17.6%<br>(15.7 – 19.6) |
| mAbs<br>+ Maternal vaccination<br>+ Older adult vaccination | 6.41<br>(5.15 – 8.22) | 0.80<br>(0.71 – 0.95) | 0.80<br>(0.71 – 0.95) | 0.15<br>(0.13 – 0.20) | 49.4%<br>(42.0 – 59.7) | 27.7%<br>(26.9 – 31.5) | 52.3%<br>(51.5 – 54.8) | 22.5%<br>(19.3 – 28.9) |
| **Thailand: the average number of RSV-associated ALRI hospitalisations averted** |  |  |  |  | **Reduction (%)** |  |  |  |
| **Intervention** | **0-11 mths** | **60-74 yrs** | **≥75 yrs** | **All ages** | **0-11 mths** | **60-74 yrs** | **≥75 yrs** | **All ages** |
| mAbs | 93<br>(71 – 128) | -0.02<br>(-0.02 – 0.19) | -0.02<br>(-0.02 – 0.19) | 0.98<br>(0.75 – 1.43) | 19.5%<br>(15.1 – 27.0) | -0.3%<br>(-0.4 – 3.5) | -0.3%<br>(-0.4 – 3.5) | 13.3%<br>(10.2 – 19.5) |
| Maternal vaccination | 120<br>(90 – 152) | -0.02<br>(-0.03 – 0.06) | -0.02<br>(-0.03 – 0.06) | 1.27<br>(0.95 – 1.62) | 25.2%<br>(19.1 – 31.7) | -0.4%<br>(-0.56 – 0.11) | -0.4%<br>(-0.55 – 0.10) | 17.1%<br>(12.9 – 22.0) |
| Older adult vaccination | 5.55<br>(4.53 – 6.60) | 1.81<br>(1.71 – 1.88) | 1.81<br>(1.71 – 1.88) | 0.52<br>(0.50 – 0.54) | 1.18%<br>(0.95 – 1.42) | 34.8%<br>(34.6 – 34.9) | 49.2%<br>(49.1 – 49.4) | 7.1%<br>(6.7 – 7.4) |
| mAbs<br>+ Maternal vaccination<br>+ Older adult vaccination | 256<br>(210 – 306) | 1.66<br>(1.56 – 1.79) | 1.66<br>(1.56 – 1.79) | 3.12<br>(2.63 – 3.70) | 53.8%<br>(44.1 – 64.6) | 31.5%<br>(31.3 – 33.9) | 37.8%<br>(37.6 – 40.0) | 42.3%<br>(35.5 – 50.8) |

For older adults, we estimated similar relative vaccine benefit among those aged 75 years compared with those aged 60–74 years. In Hong Kong, the annual proportion of RSV laboratory-confirmed cases averted was 21.7% for adults aged ≥75 years and 27.7% for those aged 60–74 years. Increasing vaccination coverage to 50% and 75% in Hong Kong further reduced severe outcomes among older adults to 55.2–61.9% and 70.3–71.6%, respectively. Similarly, in Beijing, vaccinating older adults with 30–40% coverage averted 39.7% of RSV-associated ARIs among those aged ≥75 years and 33.9% among those aged 60–74 years. In Thailand, the annual RSV-associated ALRI hospitalization rate per 100,000 population was reduced by 49.8% for adults aged ≥75 years and 34.8% for those aged 60–74 years. Combining all three interventions—long-acting mAbs, maternal vaccination, and older adult vaccination—achieved the greatest reduction in RSV severe outcomes across all age groups, reducing severe outcomes by 18.3% (14.1–27.6), 22.5% (19.3–28.9), and 42.3% (35.5–50.8) in Hong Kong, Beijing, and Thailand, respectively (**Table 2**).

## Discussion

Despite the substantial burden of RSV disease in LMICs, most studies on RSV transmission dynamics have been conducted in high-income settings, such as European nations [4, 5], Australia [6], and the United States [7], all located in temperate regions. Our study provides the first direct comparison of RSV transmission dynamics across Asian regions with markedly different seasonal patterns—specifically Beijing (temperate), Hong Kong (subtropical), and Thailand (tropical). We found that several key parameters, including the duration of immunity and relative susceptibility following reinfection, were associated with regional RSV seasonality, consistent with findings by Hodgson *et al* in the UK [4]. For example, our findings suggested that the annual infection attack rate among children under one year of age was higher in Hong Kong than in Beijing or Thailand, likely reflecting Hong Kong’s irregular seasonal pattern characterized by two annual peaks of RSV activity.

Our findings suggest that expanding access to RSV mAbs and vaccines represents an important public health strategy to reduce the burden of RSV disease among infants and older adults (**Table 2**). In Hong Kong, most laboratory-confirmed RSV cases were identified through testing prioritised for hospitalised patients with acute respiratory infections [48]. Assuming similar coverage levels for RSV vaccines and mAbs, our estimated number of averted laboratory-confirmed RSV cases among infants and older adults was comparable to hospitalization reductions reported in modelling studies from the United States by Hansen *et al* [7], and across 13 high-income countries by Du *et al* [49]. In Thailand, the estimated number of averted RSV-associated ARI hospitalizations was lower, likely due to under-ascertainment and underreporting, as RSV-related hospitalisations were only identified retrospectively from electronic health records using ICD-10 codes [15]. In Beijing, we estimated the number of averted RSV-associated ARIs reported through the RPSS surveillance network. These figures are not directly comparable to estimates from other countries because of differences in sampling and surveillance methodologies (**Figure S7**) [23].

Our findings underscore the potential for substantial health benefits from RSV interventions, such as mAbs and vaccinations, even under moderate uptake scenarios (30–40%) and conservative effectiveness assumptions, given the limited real-world data on vaccine performance beyond 180 days post-administration. However, actual uptake of RSV mAbs and vaccines will depend on factors such as public awareness, acceptability, cost, and recommendations from health authorities. Recent surveys conducted in mainland China (2023–2024) reported low awareness and perceived susceptibility to RSV infection among 42.9–55.7% of participants. These surveys also indicated that approximately 60% of respondents considered an RSV vaccine price below USD 28 acceptable [50, 51]. In contrast, the current out-of-pocket cost of RSV vaccines ranges from USD 300 to 450 in Hong Kong and USD 220 to 400 in Thailand. Furthermore, evidence on acceptance of maternal RSV vaccination remains limited, and uptake of seasonal influenza vaccination among pregnant women has been extremely low (<10%) in mainland China and Hong Kong over the past decade [52, 53].

Our study had several limitations. First, measures of severe clinical outcomes varied across the three regions: laboratory-confirmed cases in Hong Kong, RSV-associated ARIs reported to RPSS in Beijing, and RSV-associated ARI hospitalizations in Thailand (**Figure S7**). This heterogeneity may have introduced bias in parameter estimation, contributing to the substantially higher estimated infection attack rates among older adults in Beijing, and limits direct comparability of RSV transmission dynamics and intervention impacts. Second, estimation of certain parameters, such as the proportion of asymptomatic infections, relied heavily on prior assumptions, as available data were insufficient to fully inform inference (**Table S3**). Third, we calibrated each region-specific model using all available seroprevalence data, which may not accurately capture age-specific patterns given the diverse seasonality of RSV globally. Furthermore, calibration using all available seroprevalence data may not fully capture age-specific patterns in each region, particularly given rapid antibody changes in the first year of life and the influence of monthly age composition among infants. Finally, because real-world uptake data for RSV interventions in Asia are lacking, we referenced coverage estimates from Europe and the United States. Given the recent introduction of RSV vaccines, real-world experience with vaccination programs in Asia remains limited, and our assumed uptake rates may not reflect actual adoption in the three study regions. Improving awareness, accessibility, acceptability, and addressing vaccine hesitancy will be critical to increasing uptake and maximizing the impact of RSV interventions.

In conclusion, we developed an RSV transmission model integrating epidemiological data from Hong Kong, Beijing, and Thailand, and used this calibrated model to estimate the impacts of interventions, including mAbs and vaccinations. Our model provides a novel framework for evaluating intervention effectiveness, particularly in Asian populations. As real-world effectiveness data become available, future studies can leverage these insights to optimize intervention combinations for diverse settings.

## Data Availability

All data produced in the present study are available upon reasonable request to the authors.

## Ethics statement

The study was based on anonymized data that are publicly available, or aggregate data requested from the Hong Kong Centre for Health Protection. No ethical approval was required by the Institutional Review Board of the University of Hong Kong / Hospital Authority Hong Kong West Cluster (HKU / HA HKW IRB) for the use of the data (exemption obtained).

## Acknowledgement

We thank Ko Nakajo and Hiroshi Nishura (Kyoto University School of Public Health, Kyoto, Japan) for providing RSV seroprevalence data summarised in their meta-analysis [27].

## References

1. Hansen CL, Chaves SS, Demont C, Viboud C. Mortality associated with influenza and respiratory syncytial virus in the US, 1999-2018. JAMA Network Open. 2022;5(2):e220527–e.

2. Li Y, Wang X, Blau DM, Caballero MT, Feikin DR, Gill CJ, et al. Global, regional, and national disease burden estimates of acute lower respiratory infections due to respiratory syncytial virus in children younger than 5 years in 2019: a systematic analysis. The Lancet. 2022;399(10340):2047–64.

3. Shi T, McAllister DA, O’Brien KL, Simoes EAF, Madhi SA, Gessner BD, et al. Global, regional, and national disease burden estimates of acute lower respiratory infections due to respiratory syncytial virus in young children in 2015: a systematic review and modelling study. The Lancet. 2017;390(10098):946–58. doi: 10.1016/S0140-6736(17)30938-8.

4. Hodgson D, Pebody R, Panovska-Griffiths J, Baguelin M, Atkins KE. Evaluating the next generation of RSV intervention strategies: a mathematical modelling study and cost-effectiveness analysis. BMC medicine. 2020;18:1–14.

5. Krauer F, Guenther F, Treskova-Schwarzbach M, Schoenfeld V, Koltai M, Jit M, et al. Effectiveness and efficiency of immunisation strategies to prevent RSV among infants and older adults in Germany: a modelling study. BMC medicine. 2024;22(1):478.

6. Giannini F, Hogan AB, Cameron E, Le H, Minney-Smith C, Richmond P, et al. Estimating the impact of Western Australia’s first respiratory syncytial virus immunisation program for all infants: A mathematical modelling study. Vaccine. 2025;56:127155.

7. Hansen CL, Lee L, Bents SJ, Perofsky AC, Sun K, Starita LM, et al. Scenario Projections of Respiratory Syncytial Virus Hospitalizations Averted Due to New Immunizations. JAMA network open. 2025;8(6):e2514622–e.

8. Paynter S, Yakob L, Simoes EA, Lucero MG, Tallo V, Nohynek H, et al. Using mathematical transmission modelling to investigate drivers of respiratory syncytial virus seasonality in children in the Philippines. PLoS One. 2014;9(2):e90094.

9. Shaaban FL, Groenendijk RW, Baral R, Caballero MT, Crowe JE, Jr., Englund JA, et al. The path to equitable respiratory syncytial virus prevention for infants: challenges and opportunities for global implementation. The Lancet Global Health. doi: 10.1016/S2214-109X(25)00379-1.

10. Broor S, Campbell H, Hirve S, Hague S, Jackson S, Moen A, et al. Leveraging the Global Influenza Surveillance and Response System for global respiratory syncytial virus surveillance—opportunities and challenges. Influenza and other respiratory viruses. 2020;14(6):622–9.

11. Zhang Y, Yuan L, Zhang Y, Zhang X, Zheng M, Kyaw MH. Burden of respiratory syncytial virus infections in China: systematic review and meta–analysis. Journal of global health. 2015;5(2).

12. Centre for Health Protection of The Government of the Hong Kong Special Administrative Region. RSV disease and Interim Consensus on the use of RSV vaccines in Hong Kong 2024. Available from: https://www.chp.gov.hk/files/pdf/cdw_v21_1.pdf.

13. Chan PK, Tam WW, Lee TC, Hon KL, Lee N, Chan MC, et al. Hospitalization incidence, mortality, and seasonality of common respiratory viruses over a period of 15 years in a developed subtropical city. Medicine. 2015;94(46).

14. Naorat S, Chittaganpitch M, Thamthitiwat S, Henchaichon S, Sawatwong P, Srisaengchai P, et al. Hospitalizations for acute lower respiratory tract infection due to respiratory syncytial virus in Thailand, 2008–2011. The Journal of infectious diseases. 2013;208(suppl_3):S238–S45.

15. Sitthikarnkha P, Uppala R, Niamsanit S, Sutra S, Thepsuthammarat K, Techasatian L, et al. Burden of Respiratory Syncytial Virus Related Acute Lower Respiratory Tract Infection in Hospitalized Thai Children: A 6-Year National Data Analysis. Children. 2022;9(12):1990.

16. Mazur NI, Terstappen J, Baral R, Bardají A, Beutels P, Buchholz UJ, et al. Respiratory syncytial virus prevention within reach: The vaccine and monoclonal antibody landscape. The Lancet Infectious Diseases. 2022.

17. Drysdale SB, Cathie K, Flamein F, Knuf M, Collins AM, Hill HC, et al. Nirsevimab for prevention of hospitalizations due to RSV in infants. New England Journal of Medicine. 2023;389(26):2425–35.

18. Munro AP, Drysdale SB, Cathie K, Flamein F, Knuf M, Collins AM, et al. 180-day efficacy of nirsevimab against hospitalisation for respiratory syncytial virus lower respiratory tract infections in infants (HARMONIE): a randomised, controlled, phase 3b trial. The Lancet Child & Adolescent Health. 2025;9(6):404–12.

19. Kelleher K, Subramaniam N, Drysdale SB. The recent landscape of RSV vaccine research. Therapeutic Advances in Vaccines and Immunotherapy. 2025;13:25151355241310601.

20. Bajema KL, Yan L, Li Y, Argraves S, Rajeevan N, Fox A, et al. Respiratory syncytial virus vaccine effectiveness among US veterans, September, 2023 to March, 2024: a target trial emulation study. The Lancet Infectious Diseases. 2025.

21. Chiu SS, Cowling BJ, Peiris JM, Chan EL, Wong WH, Lee KP. Effects of nonpharmaceutical COVID-19 interventions on pediatric hospitalizations for other respiratory virus infections, Hong Kong. Emerging Infectious Diseases. 2022;28(1):62.

22. Lau YC, Ryu S, Du Z, Wang L, Wu P, Lau EH, et al. Impact of COVID-19 control measures on respiratory syncytial virus and hand-foot-and-mouth disease transmission in Hong Kong and South Korea. Epidemics. 2024;49:100797.

23. Li M, Cong B, Wei X, Wang Y, Kang L, Gong C, et al. Characterising the changes in RSV epidemiology in Beijing, China during 2015–2023: results from a prospective, multi-centre, hospital-based surveillance and serology study. The Lancet Regional Health–Western Pacific. 2024;45.

24. Lu G, Gonzalez R, Guo L, Wu C, Wu J, Vernet G, et al. Large-scale seroprevalence analysis of human metapneumovirus and human respiratory syncytial virus infections in Beijing, China. Virology journal. 2011;8(1):1–10.

25. Pasittungkul S, Thongpan I, Vichaiwattana P, Thongmee T, Klinfueng S, Suntronwong N, et al. High seroprevalence of antibodies against human respiratory syncytial virus and evidence of respiratory syncytial virus reinfection in young children in Thailand. International Journal of Infectious Diseases. 2022;125:177–83.

26. Cohen CA, Grzelak L, Chiu SS, Hui DS, Kwan MY, Tsang OT, et al. SARS-CoV-2 antibody responses in children exhibit higher FcR engagement and avidity than in adults. Nature Communications. 2025;16(1):7879.

27. Nakajo K, Nishiura H. Age-dependent risk of respiratory syncytial virus infection: A systematic review and hazard modeling from serological data. The Journal of Infectious Diseases. 2023;228(10):1400–9.

28. Bloom-Feshbach K, Alonso WJ, Charu V, Tamerius J, Simonsen L, Miller MA, et al. Latitudinal variations in seasonal activity of influenza and respiratory syncytial virus (RSV): a global comparative review. PloS one. 2013;8(2):e54445.

29. Leung K, Jit M, Lau EH, Wu JT. Social contact patterns relevant to the spread of respiratory infectious diseases in Hong Kong. Scientific reports. 2017;7(1):7974.

30. Mistry D, Litvinova M, Pastore y Piontti A, Chinazzi M, Fumanelli L, Gomes MF, et al. Inferring high-resolution human mixing patterns for disease modeling. Nature communications. 2021;12(1):323.

31. Prem K, Zandvoort Kv, Klepac P, Eggo RM, Davies NG, Group CftMMoIDC-W, et al. Projecting contact matrices in 177 geographical regions: an update and comparison with empirical data for the COVID-19 era. PLoS computational biology. 2021;17(7):e1009098.

32. Pecenka C, Sparrow E, Feikin DR, Srikantiah P, Darko DM, Karikari-Boateng E, et al. Respiratory syncytial virus vaccination and immunoprophylaxis: realising the potential for protection of young children. The Lancet. 2024;404(10458):1157–70.

33. Hon K, Cheung EW, Li AM, Fung GP, Lam DS, Lee MS, et al. Practice recommendations for respiratory syncytial virus prophylaxis among children in Hong Kong. Hong Kong Medical Journal. 2025;31(1):48.

34. Deng K, Liang J, Mu Y, Liu Z, Wang Y, Li M, et al. Preterm births in China between 2012 and 2018: an observational study of more than 9 million women. The Lancet Global Health. 2021;9(9):e1226–e41.

35. Wang X, Li Y, Shi T, Bont LJ, Chu HY, Zar HJ, et al. Global disease burden of and risk factors for acute lower respiratory infections caused by respiratory syncytial virus in preterm infants and young children in 2019: a systematic review and meta-analysis of aggregated and individual participant data. The Lancet. 2024;403(10433):1241–53.

36. Hammitt LL, Dagan R, Yuan Y, Baca Cots M, Bosheva M, Madhi SA, et al. Nirsevimab for prevention of RSV in healthy late-preterm and term infants. New England Journal of Medicine. 2022;386(9):837–46.

37. Centers for Disease Control Prevention. Respiratory syncytial virus (RSV) vaccination coverage, pregnant persons, United States. 2024.

38. Kampmann B, Madhi SA, Munjal I, Simões EAF, Pahud BA, Llapur C, et al. Bivalent Prefusion F Vaccine in Pregnancy to Prevent RSV Illness in Infants. New England Journal of Medicine. 2023;388(16):1451–64. doi: doi:10.1056/NEJMoa2216480.

39. Kriss JL. Influenza, COVID-19, and Respiratory Syncytial Virus Vaccination Coverage Among Adults—United States, Fall 2024. MMWR Morbidity and Mortality Weekly Report. 2024;73.

40. Centre for Health Protection of The Government of the Hong Kong Special Administrative Region. Statistics on vaccination programmes in the past 3 years 2025. Available from: https://www.chp.gov.hk/en/features/102226.html.

41. Centers for Disease Control Prevention. Influenza Vaccination Coverage and Intent for Vaccination, Adults 18 Years and Older, United States 2025. Available from: https://www.cdc.gov/fluvaxview/dashboard/adult-coverage.html.

42. Shen Y, Wang J, Lv M, Wu J, Nicholas S, Maitland E, et al. Predicting future vaccination habits: The link between influenza vaccination patterns and future vaccination decisions among old aged adults in China. Journal of Infection and Public Health. 2024;17(6):1079–85.

43. Montgomery MP, Praphasiri P, Ditsungnoen D, Akarasewi P, Chittaganpitch M, Puthavathana P, et al. Influenza surveillance and vaccine policy in Thailand—a historical perspective. The Lancet Regional Health-Southeast Asia. 2025;41.

44. Payne AB, Watts JA, Mitchell PK, Dascomb K, Irving SA, Klein NP, et al. Respiratory syncytial virus (RSV) vaccine effectiveness against RSV-associated hospitalisations and emergency department encounters among adults aged 60 years and older in the USA, October, 2023, to March, 2024: a test-negative design analysis. The Lancet. 2024;404(10462):1547–59. doi: 10.1016/S0140-6736(24)01738-0.

45. Liu D, Leung K, Jit M, Wu JT. Cost-effectiveness of strategies for preventing paediatric lower respiratory infections associated with respiratory syncytial virus in eight Chinese cities. Vaccine. 2021;39(39):5490–8. doi: 10.1016/j.vaccine.2021.08.057.

46. Hall CB, Walsh EE, Long CE, Schnabel KC. Immunity to and frequency of reinfection with respiratory syncytial virus. Journal of Infectious Diseases. 1991;163(4):693–8.

47. Yang B, Lin Y, Xiong W, Liu C, Gao H, Ho F, et al. Comparison of control and transmission of COVID-19 across epidemic waves in Hong Kong: an observational study. The Lancet Regional Health–Western Pacific. 2024;43.

48. Centre for Health Protection of The Government of the Hong Kong Special Administrative Region. Detection of pathogens from respiratory specimens 2025. Available from: https://www.chp.gov.hk/en/statistics/data/10/641/642/2274.html.

49. Du Z, Pandey A, Moghadas SM, Bai Y, Wang L, Matrajt L, et al. Impact of RSVpreF vaccination on reducing the burden of respiratory syncytial virus in infants and older adults. Nature Medicine. 2025:1–6.

50. Wang Q, Xiu S, Yang L, Li L, Yang M, Wang X, et al. Perceptions about respiratory syncytial virus (RSV) and attitudes toward the RSV vaccine among the general public in China: A cross-sectional survey. Human Vaccines & Immunotherapeutics. 2024;20(1):2310916.

51. Wang Q, Yang L, Li L, Xiu S, Yang M, Wang X, et al. Investigating parental perceptions of respiratory syncytial virus (RSV) and attitudes to RSV vaccine in Jiangsu, China: insights from a cross-section study. Vaccine. 2025;44:126570.

52. Yuen CYS, Fong DYT, Lee ILY, Chu S, Siu ES-m, Tarrant M. Prevalence and predictors of maternal seasonal influenza vaccination in Hong Kong. Vaccine. 2013;31(45):5281–8.

53. Li R, Xie R, Yang C, Rainey J, Song Y, Greene C. Identifying ways to increase seasonal influenza vaccine uptake among pregnant women in China: A qualitative investigation of pregnant women and their obstetricians. Vaccine. 2018;36(23):3315–22.

