## Supplementary information for "Estimating the impact of vaccination and long-acting monoclonal antibodies for RSV epidemics across Hong Kong, Beijing, and Thailand: a modelling study"

**RSV transmission model**

***Model structure***

We developed an age-structure compartmental RSV transmission model adapted from Hodgson *et al* (see **Figure S1** model diagram and **Table S1** for the description of epidemiological state variables of the model) [1]. In the absence of RSV mAbs and vaccines, the model assumes that infants become completely susceptible (in the compartment $S_0$) soon after being partially protected against infection by maternally derived immunity for a short time. Individuals experience frequent reinfections throughout life, where infectivity and duration of infectiousness depend on the number of prior infections (in the compartments $I_0$, $I_1$, $I_2$ and $I_3$), and susceptibility is gradually reduced with the build-up of partial immunity through repeated infections (in the compartments $S_1$, $S_2$ and $S_3$). The model also assumes sterilising immunity by including recovered compartments following each infection ($R_0$, $R_1$, $R_2$ and $R_3$).

Specifically, in the transmission model, we assumed infants are born into either the $M$ compartment with complete protection against infection or the $S_0$ compartment with no protection (**Figure S1**). The proportion born into $S_0$ is equivalent to that of adults aged 25-44 who are estimated to be susceptible (i.e., $1 - p_m(t)$, see explanation in the section "***Maternal protection***" below). After maternal protection wanes, infants in the $M$ compartment become fully susceptible ($S_0$). Following the first infection ($I_0$), individuals have a short period on sterilising immunity ($R_0$). After this immunity wanes, individuals are susceptible again but with reduced susceptibility ($S_1$). Following each infection the duration of infectiousness becomes shorter, the relative infectivity decreases, and the proportion of asymptomatic infections increases.

To capture the heterogeneity in transmission, we stratified the model by age (indicated by superscript $a$), and all compartments are broken down by 25 age groups, which allows for age-specific demographic and contact data to be used and RSV infections and interventions among infants to be closely monitored (including age groups <1, 1, 2, 3, 4, 5, 6, 7, 8, 9, 10, 11 months, and 1, 2, 3, 4, 5-9, 10-14, 15-24, 25-34, 35-44, 45-54, 55-64, 65-74, ≥75 years). The differential equations describing the transmission dynamics of RSV are as follows, with $a$ denoting the age group:

$$\frac{dM^a}{dt} = p_m(t)\mu[\![a = 1]\!] - \xi M^a - \eta^a M^a + \eta^{a-1} M^{a-1}$$

$$\frac{dS_0^a}{dt} = (1 - p_m(t))\mu[\![a = 1]\!] + \xi M^a - \lambda_0^a(t)S_0^a - \eta^a S_0^a + \eta^{a-1}S_0^{a-1}$$

$$\frac{dE_0^a}{dt} = \lambda_0^a(t)S_0^a - \sigma E_0^a - \eta^a E_0^a + \eta^{a-1}E_0^{a-1}$$

$$\frac{dA_0^a}{dt} = p_{asym}^a \sigma E_0^a - \gamma_0 A_0^a - \eta^a A_0^a + \eta^{a-1}A_0^{a-1}$$

$$\frac{dI_0^a}{dt} = \left(1 - p_{asym}^a\right)\sigma E_0^a - \gamma_0 I_0^a - \eta^a I_0^a + \eta^{a-1}I_0^{a-1}$$

$$\frac{dR_0^a}{dt} = \gamma_0 A_0^a + \gamma_0 I_0^a - \omega R_0^a - \eta^a R_0^a + \eta^{a-1}R_0^{a-1}$$

$$\frac{dS_1^a}{dt} = \omega R_0^a - \lambda_1^a(t)S_1^a - \eta^a S_1^a + \eta^{a-1}S_1^{a-1}$$

$$\frac{dE_1^a}{dt} = \lambda_1^a(t)S_1^a - \sigma E_1^a - \eta^a E_1^a + \eta^{a-1}E_1^{a-1}$$

$$\frac{dA_1^a}{dt} = p_{asym}^a \sigma E_1^a - \gamma_1 A_1^a - \eta^a A_1^a + \eta^{a-1}A_1^{a-1}$$

$$\frac{dI_1^a}{dt} = \left(1 - p_{asym}^a\right)\sigma E_1^a - \gamma_1 I_1^a - \eta^a I_1^a + \eta^{a-1}I_1^{a-1}$$

$$\frac{dR_1^a}{dt} = \gamma_1 A_1^a + \gamma_1 I_1^a - \omega R_1^a - \eta^a R_1^a + \eta^{a-1}R_1^{a-1}$$

$$\frac{dS_2^a}{dt} = \omega R_1^a - \lambda_2^a(t)S_2^a - \eta^a S_2^a + \eta^{a-1}S_2^{a-1}$$

$$\frac{dE_2^a}{dt} = \lambda_2^a(t)S_2^a - \sigma E_2^a - \eta^a E_2^a + \eta^{a-1}E_2^{a-1}$$

$$\frac{dA_2^a}{dt} = p_{asym}^a \sigma E_2^a - \gamma_2 A_2^a - \eta^a A_2^a + \eta^{a-1}A_2^{a-1}$$

$$\frac{dI_2^a}{dt} = \left(1 - p_{asym}^a\right)\sigma E_2^a - \gamma_2 I_2^a - \eta^a I_2^a + \eta^{a-1}I_2^{a-1}$$

$$\frac{dR_2^a}{dt} = \gamma_2 A_2^a + \gamma_2 I_2^a - \omega R_2^a - \eta^a R_2^a + \eta^{a-1}R_2^{a-1}$$

$$\frac{dS_3^a}{dt} = \omega R_2^a + \omega R_3^a - \lambda_3^a(t)S_3^a - \eta^a S_3^a + \eta^{a-1}S_3^{a-1}$$

$$\frac{dE_3^a}{dt} = \lambda_3^a(t)S_3^a - \sigma E_3^a - \eta^a E_3^a + \eta^{a-1}E_3^{a-1}$$

$$\frac{dA_3^a}{dt} = p_{asym}^a \sigma E_3^a - \gamma_3 A_3^a - \eta^a A_3^a + \eta^{a-1}A_3^{a-1}$$

$$\frac{dI_3^a}{dt} = \left(1 - p_{asym}^a\right)\sigma E_3^a - \gamma_3 I_3^a - \eta^a I_3^a + \eta^{a-1}I_3^{a-1}$$

$$\frac{dR_3^a}{dt} = \gamma_3 A_3^a + \gamma_3 I_3^a - \omega R_3^a - \eta^a R_3^a + \eta^{a-1}R_3^{a-1}$$

$$\frac{dX^a}{dt} = \sigma(E_0^a + E_1^a + E_2^a + E_3^a)$$

where $[\![a=1]\!]$ is the indicator function. $p_m(t)$ is the proportion of newborn infants who are completely protected by maternally derived immunity (see the formulation as follows). The duration of secondary and subsequent infections was shortened and thus $\gamma_1 = \gamma_0/g_1$ and $\gamma_2 = \gamma_3 = \gamma_0/g_1/g_2$.

#### *Parameterisation with fixed demographic and contact data*

We considered a population with natural births and deaths to simulation the RSV transmission dynamics in the model. To maintain the population size of each age group, we assumed a pro-rata immigration rate of each age group (to all compartments $S_i^a$, $E_i^a$, $A_i^a$, $I_i^a$, $R_i^a$ except $M^a$) to adjust for the difference between birth rate $\mu$ and aging/death rate of $\eta^{\geq 75}$. We extracted age-specific population sizes and birth rates for Beijing, Hong Kong and Thailand from the respective census department for the year 2024 [2-4]. The annual birth rates were divided by 365.25 to convert to an average daily number of new births, and the rate of aging was assumed to be the inverse of the time spent in each age class. We obtained the contact data to describe the daily rate of physical and conversational contacts made by age group $a$ with age group $b$ from Mistry *et al* for Beijing, Leung *et al* for Hong Kong, and Prem *et al* for Thailand [5-7].

#### *Maternal protection*

We assumed that maternal protection is mediated through maternal antibodies. Since all compartments are stratified into 25 age groups, we modelled maternal protection, denoted as $p_m(t)$ at time $t$ under the following assumptions:

1) Pregnant women in compartments $E_i^{a'}$, $A_i^{a'}$, $I_i^{a'}$ do not provide any maternal protection.
2) Pregnant women in compartment $M^{a'}$ provide full maternal protection.
3) Pregnant women in compartments $R_i^{a'}$ and $S_i^{a'}$ provide partial protection adjusted by the proportion of seropositivity $\pi$ and reduced susceptibility to repeat infections $\left(1-\prod_{k=0}^{i}\delta_k\right)$.

Therefore, we modelled the dynamics of $p_m(t)$ as follows:

$$p_m(t) = \frac{\sum_{a'}\left(M^{a'}(t) + \pi\left(\sum_{i=0}^{3} R_i^{a'}(t) + \sum_{i=0}^{3}\left(1-\prod_{k=0}^{i}\delta_k\right)S_i^{a'}(t)\right)\right)}{\sum_{a'} N^{a'}}$$

where $a'$ indicates the child-bearing age groups (i.e., assuming to be 25-44 years in Hong Kong, Beijing and Thailand), $N^{a'}$ is the population size of the age group $a'$, and $\pi$ represents the probability of IgG ELISA seropositivity immediately after RSV infection. $\delta_0 = 1$ is the susceptibility to primary infection, and $\delta_1$, $\delta_2$ and $\delta_3$ are relative susceptibility to secondary, tertiary and subsequent infections.

***Seasonality and force of infection***

Following Hodgson *et al* [1], we assumed that the probability of RSV transmission for a contact made between two age groups is $q_p$ if the contact is physical and $q_p q_c$ if the contact is conversational, where $0 < 1 - q_c < 1$ is the relative reduction in transmission. Similarly, we assumed that the infectiousness of asymptomatic infections is reduced, and the relative infectiousness is $0 < \alpha < 1$. Finally, the probability of transmission is seasonally forced according to a normal distribution, with $b_1$ and $\varphi$ represent the amplitude and timing of the seasonal RSV peak transmission, and $\psi$ represents the standard deviation. Thus, the force of infection is formulated as follows:

$$\lambda_i^a(t) =$$
$$q_p\left(1 + b_1 \exp\left((mod(t, 365)/365 - \varphi)^2/(2\psi^2)\right)\right)\prod_{k=0}^{i}\delta_k\sum_{b=1}^{25}\frac{\left(c_{phy}^{a,b} + q_c c_{con}^{a,b}\right)}{N^b}\left(\alpha A_i^b + I_i^b\right)$$

where $N^b$ is the population size of the age group $b$.

***Model output***

The output of the epidemic model is the weekly or monthly number of RSV infections:

$$x_{w_k}^a = X^a\left(t_{w_k}\right) - X^a\left(t_{w_{k-1}}\right)$$

$$x_{m_k}^a = X^a\left(t_{m_k}\right) - X^a\left(t_{m_{k-1}}\right)$$

where $X^a\left(t_{w_{k-1}}\right)$ and $X^a\left(t_{w_k}\right)$ are the cumulative number of RSV infections by week $w_{k-1}$ and $w_k$, and $X^a\left(t_{m_{k-1}}\right)$ and $X^a\left(t_{m_k}\right)$ are the cumulative number of RSV infections by month $m_{k-1}$ and $m_k$, respectively. Similarly, the age-specific seroprevalence is

$$p_s^a(t) = \frac{M^a(t) + \pi\left(\sum_{i=0}^{3} R_i^a(t) + \sum_{i=0}^{3}\left(1 - \prod_{k=0}^{i}\delta_k\right)S_i^a(t)\right)}{N^a}$$

where $p_s^a(t)$ is the seropositivity proportion of age group $a$ measured by RSV IgG ELISA at time $t$.

**Modelling RSV interventions with mAbs and vaccinations**

To evaluate the impact of intervention programs involving mAbs and vaccinations, we stratified newborn infants into separate compartments (**Table S1**): those receiving and protected by long-acting mAbs at birth ($V_{mAb1}$, $V_{mAb2}$), those covered and protected by maternal vaccination ($V_{mvax1}$, $V_{mvax2}$), those protected by maternally derived immunity ($M$) and those completely susceptible ($S_0$) (**Figure S2**). For mAbs and maternal vaccination, we assumed an "all-or-nothing" protection mechanism and applied an Erlang distribution by splitting immunized infants into two successive compartments [8]. Specifically, given $p_{mAb}$, the effectiveness of mAb protection within $d_{mAb} = 180$ days, and the shape parameter $\alpha_{mAb} = 2$, we estimated the scale parameter $\beta_{mAb}$ as:

$$\beta_{mAb} = \frac{d_{mAb}\,\Gamma(\alpha_{mAb})}{\Gamma\left(\alpha_{mAb}, \frac{1}{1 - p_{mAb}}\right)}$$

And the parameter for mAb protection duration was estimated as:

$$\xi_{mAb} = \frac{1}{\beta_{mAb}}$$

Similarly, we estimated the parameter for protection duration of maternal vaccination as:

$$\xi_{mvax} = \frac{\Gamma\left(\alpha_{mvax}, \frac{1}{1 - p_{mvax}}\right)}{d_{mvax}\,\Gamma(\alpha_{mvax})}$$

For vaccinating older adults, we assumed only individuals aged 60 or above in the $S_3$ compartments were eligible for vaccination, which confers full protection against infection (**Figure S2**). Vaccinated older adults transition from $S_3$ into compartments $V_{pvax1}$ compartments at a constant rate $\gamma_{pvax}$, with vaccine uptake assumed to be achieved within 180 days after entering the age group (**Table S2**). Subsequently, individuals move from $V_{pvax1}$ to $V_{pvax2}$, and then return from $V_{pvax2}$ to $S_3$, at a constant rate of $\xi_{pvax}$, provided they are not infected. We assumed an "all-or-nothing" vaccine and individuals are fully protected from infection when they are in the compartments $V_{pvax1}$ and $V_{pvax2}$, and the estimated the duration of protection accounting for vaccine effectiveness $p_{pvax}$:

$$\xi_{pvax} = \frac{\Gamma\left(\alpha_{pvax}, \frac{1}{1 - p_{pvax}}\right)}{d_{pvax}\,\Gamma(\alpha_{pvax})}$$

The differential equations of the RSV transmission dynamics with mAb and vaccination are as follows:

$$\frac{dV_{mvax1}^{a}}{dt} = c_{mvax}\mu[\![a=1]\!] - \xi_{mvax}V_{mvax1}^{a} - \eta^{a}V_{mvax1}^{a} + \eta^{a-1}V_{mvax1}^{a-1}$$

$$\frac{dV_{mvax2}^{a}}{dt} = \xi_{mvax}V_{mvax1}^{a} - \xi_{mvax}V_{mvax2}^{a} - \eta^{a}V_{mvax2}^{a} + \eta^{a-1}V_{mvax2}^{a-1}$$

$$\frac{dV_{mAb1}^{a}}{dt} = c_{mAb}\mu[\![a=1]\!] - \xi_{mAb}V_{mAb1}^{a} - \eta^{a}V_{mAb1}^{a} + \eta^{a-1}V_{mAb1}^{a-1}$$

$$\frac{dV_{mAb2}^{a}}{dt} = \xi_{mAb}V_{mAb1}^{a} - \xi_{mAb}V_{mAb2}^{a} - \eta^{a}V_{mAb2}^{a} + \eta^{a-1}V_{mAb2}^{a-1}$$

$$\frac{dM^{a}}{dt} = (1 - c_{mAb} - c_{mvax})p_{m}(t)\mu[\![a=1]\!] - \xi M^{a} - \eta^{a}M^{a} + \eta^{a-1}M^{a-1}$$

$$\frac{dS_{0}^{a}}{dt} = (1 - c_{mAb} - c_{mvax})(1 - p_{m}(t))\mu[\![a=1]\!] + \xi M^{a} + \xi_{mvax}V_{mvax2}^{a} + \xi_{mAb}V_{mAb2}^{a} - \lambda_{0}^{a}(t)S_{0}^{a} - \eta^{a}S_{0}^{a} + \eta^{a-1}S_{0}^{a-1}$$

$$\frac{dE_{0}^{a}}{dt} = \lambda_{0}^{a}(t)S_{0}^{a} - \sigma E_{0}^{a} - \eta^{a}E_{0}^{a} + \eta^{a-1}E_{0}^{a-1}$$

$$\frac{dA_{0}^{a}}{dt} = p_{asym}^{a}\sigma E_{0}^{a} - \gamma_{0}A_{0}^{a} - \eta^{a}A_{0}^{a} + \eta^{a-1}A_{0}^{a-1}$$

$$\frac{dI_{0}^{a}}{dt} = \left(1 - p_{asym}^{a}\right)\sigma E_{0}^{a} - \gamma_{0}I_{0}^{a} - \eta^{a}I_{0}^{a} + \eta^{a-1}I_{0}^{a-1}$$

$$\frac{dR_{0}^{a}}{dt} = \gamma_{0}A_{0}^{a} + \gamma_{0}I_{0}^{a} - \omega R_{0}^{a} - \eta^{a}R_{0}^{a} + \eta^{a-1}R_{0}^{a-1}$$

$$\frac{dS_{1}^{a}}{dt} = \omega R_{0}^{a} - \lambda_{1}^{a}(t)S_{1}^{a} - \eta^{a}S_{1}^{a} + \eta^{a-1}S_{1}^{a-1}$$

$$\frac{dE_{1}^{a}}{dt} = \lambda_{1}^{a}(t)S_{1}^{a} - \sigma E_{1}^{a} - \eta^{a}E_{1}^{a} + \eta^{a-1}E_{1}^{a-1}$$

$$\frac{dA_{1}^{a}}{dt} = p_{asym}^{a}\sigma E_{1}^{a} - \gamma_{1}A_{1}^{a} - \eta^{a}A_{1}^{a} + \eta^{a-1}A_{1}^{a-1}$$

$$\frac{dI_{1}^{a}}{dt} = \left(1 - p_{asym}^{a}\right)\sigma E_{1}^{a} - \gamma_{1}I_{1}^{a} - \eta^{a}I_{1}^{a} + \eta^{a-1}I_{1}^{a-1}$$

$$\frac{dR_{1}^{a}}{dt} = \gamma_{1}A_{1}^{a} + \gamma_{1}I_{1}^{a} - \omega R_{1}^{a} - \eta^{a}R_{1}^{a} + \eta^{a-1}R_{1}^{a-1}$$

$$\frac{dS_{2}^{a}}{dt} = \omega R_{1}^{a} - \lambda_{2}^{a}(t)S_{2}^{a} - \eta^{a}S_{2}^{a} + \eta^{a-1}S_{2}^{a-1}$$

$$\frac{dE_{2}^{a}}{dt} = \lambda_{2}^{a}(t)S_{2}^{a} - \sigma E_{2}^{a} - \eta^{a}E_{2}^{a} + \eta^{a-1}E_{2}^{a-1}$$

$$\frac{dA_{2}^{a}}{dt} = p_{asym}^{a}\sigma E_{2}^{a} - \gamma_{2}A_{2}^{a} - \eta^{a}A_{2}^{a} + \eta^{a-1}A_{2}^{a-1}$$

$$\frac{dI_{2}^{a}}{dt} = \left(1 - p_{asym}^{a}\right)\sigma E_{2}^{a} - \gamma_{2}I_{2}^{a} - \eta^{a}I_{2}^{a} + \eta^{a-1}I_{2}^{a-1}$$

$$\frac{dR_2^a}{dt} = \gamma_2 A_2^a + \gamma_2 I_2^a - \omega R_2^a - \eta^a R_2^a + \eta^{a-1} R_2^{a-1}$$

$$\frac{dS_3^a}{dt} = \omega R_2^a + \omega R_3^a - \gamma_{pvax} S_3^a + \xi_{pvax} V_{pvax2}^a - \lambda_3^a(t) S_3^a - \eta^a S_3^a + \eta^{a-1} S_3^{a-1}$$

$$\frac{dV_{pvax1}^a}{dt} = \gamma_{pvax} S_3^a - \xi_{pvax} V_{pvax1}^a - \eta^a V_{pvax1}^a + \eta^{a-1} V_{pvax1}^{a-1}$$

$$\frac{dV_{pvax2}^a}{dt} = \xi_{pvax} V_{pvax1}^a - \xi_{pvax} V_{pvax2}^a - \eta^a V_{pvax2}^a + \eta^{a-1} V_{pvax2}^{a-1}$$

$$\frac{dE_3^a}{dt} = \lambda_3^a(t) S_3^a - \sigma E_3^a - \eta^a E_3^a + \eta^{a-1} E_3^{a-1}$$

$$\frac{dA_3^a}{dt} = p_{asym}^a \sigma E_3^a - \gamma_3 A_3^a - \eta^a A_3^a + \eta^{a-1} A_3^{a-1}$$

$$\frac{dI_3^a}{dt} = \left(1 - p_{asym}^a\right) \sigma E_3^a - \gamma_3 I_3^a - \eta^a I_3^a + \eta^{a-1} I_3^{a-1}$$

$$\frac{dR_3^a}{dt} = \gamma_3 A_3^a + \gamma_3 I_3^a - \omega R_3^a - \eta^a R_3^a + \eta^{a-1} R_3^{a-1}$$

$$\frac{dX^a}{dt} = \sigma(E_0^a + E_1^a + E_2^a + E_3^a)$$

***Coverage, uptake and effectiveness of RSV interventions***

*Baseline scenario.* Before 2022, none of the three regions had a consensus on using mAb for RSV prophylaxis [9]. Although Hong Kong experts reached agreement in July 2022 to consider a 6-month regimen of mAb prophylaxis for preterm infants (<29 weeks' gestational age) and high-risk infants under the age of 1 year [10], no formal regional recommendations for RSV prophylaxis using mAbs or vaccines had been adopted in any of the three settings by the end of 2024. For the purposes of this study, we assumed no administration of RSV mAbs or vaccines across any age group in any region during the baseline period (2014–2019). This baseline scenario serves as the reference point against which we evaluate the projected public health impact of alternative intervention strategies.

*Long acting mAb use.* Based on an observational study of over 9 million Chinese women (2012-2018), we assumed that 6.4% of infants born preterm would receive long acting mAb (e.g., nirsevimab and clesrovimab) [11]. Following estimates by Wang *et al.*, we assumed that preterm infants accounted for 25% (95% uncertainty range: 16–37) of severe outcomes (e.g., hospitalizations) due to RSV-associated acute lower respiratory infection (ALRI) among all infants, regardless of gestational age [12]. Based on results from the recent MELODY trial, we assumed the long-acting mAb provided 77.3% (95% CI: 50.3 – 89.7) efficacy against RSV-associated hospitalizations for an average duration of 180 days post-administration [13]. An earlier randomized controlled trial (RCT) conducted in 2016-2017 among preterm infants only reported a similar efficacy of 78.4% (95% CI: 51.9–90.3) [14].

*Maternal vaccination.* We assumed that 38.5% (95% CI: 23.2 – 53.5) of pregnant women would receive maternal vaccination, based on early 2025 data from the U.S. CDC [15]. For these vaccinated mothers, we parameterised the model using Abrysvo's efficacy against severe lower respiratory tract infection (LRTI), estimated at 69.4% (95% CI: 44.3–84.1) for an average duration of 180 days after birth (**Table S2**) [16]. When both long-acting mAbs and maternal vaccination were considered, we assumed that preterm infants would receive either long-acting mAbs or be covered by maternal vaccination through their mothers.

*Vaccinating older adults*. As subsidized RSV vaccination is currently unavailable for older adults in the three regions, we assumed RSV vaccination coverage of 39.7% (95% CI: 23.1 – 58.9) for individuals aged 75 or above and 31.4% (95% CI: 18.5 – 45.7) for those aged 60-74, based on the recent US CDC data [17]. For reference, seasonal influenza vaccine uptake in Hong Kong during 2024–2025 was 25.6% among individuals aged 50–64 years and 51.8% among those aged ≥65 years [18], compared with 48.4% and 71.8%, respectively, in the United States [19]. In Thailand, coverage among older adults was approximately 47% in 2024, whereas a recent study reported substantially lower influenza vaccine uptake among older adults in Beijing: 63% sporadically vaccinated, 19% occasionally vaccinated, and 18% frequently vaccinated [20, 21]. We parameterized the model for protection against adult hospitalizations using real-world vaccine effectiveness estimates of 80% (95% CI: 71–85) from the U.S. during the 2023–2024 season [22], and assumed the same efficacy for protection against infection (**Table S2**).

**Model calibration and parameter estimation**

***Calibration to data between 2014 and 2019***

Although RSV epidemiology exhibits geographic and temporal heterogeneity with distinct seasonality [23], most seroprevalence studies demonstrate similar patterns: seroprevalence is lowest among infants aged 0–1 year, increases during early childhood, and reaches a plateau in older children and adults [24]. Accordingly, for each of the three regions, we fit the RSV transmission model to region-specific hospital admission or laboratory-confirmed case data while integrating all available seroprevalence data into the calibration. To minimize the confounding effects of the COVID-19 pandemic, our primary model calibration focused on the period from 2014 to 2019. For Hong Kong, we additionally incorporated laboratory-confirmed

RSV case data from January to March 2020 and from June 2021 to December 2024 to inform estimates of the impact of PHSMs on RSV dynamics [25].

We fit the model in two steps. First, we followed the model initialisation approach described in Hodgson *et al* [1], but did not fix $I_1(0)$ and $I_2(0)$ (**Table S2**). The model was then run for over 10 years, starting from May 1, 2004, as a burn-in period. After this, we calibrated the model to age-stratified hospital admission or laboratory-confirmed case data, which were available from July 2015 for Beijing, January 2014 for Hong Kong, and October 2014 for Thailand.

***Calibration to Hong Kong data between 2020 and 2024***

Post-pandemic age-stratified laboratory-confirmed case data were available only for Hong Kong. Therefore, we divided model calibration into two periods: pre-pandemic (2014–2019) and post-pandemic (2020–2024). We assumed that the impacts of PHSMs implemented during the COVID-19 pandemic were temporary and did not alter the intrinsic RSV transmission dynamics or severity [26]. The fitted model, initially calibrated using 2014–2019 data, was then refit to post-pandemic age-stratified case data to estimate the effects of PHSMs (i.e., estimating $\kappa_{a,1}, \ldots, \kappa_{a,5}$ only with posterior mean of other parameters in **Table S2**).

We assumed that PHSM implementation reduced, and PHSM relaxation increased, social contact patterns affecting RSV transmission. We applied linear interpolation to model the associated parameters as follows:

$$\kappa_a(t) = \begin{cases} 1 + \frac{(\kappa_{a,1}-1)(t-t_{PHSM,1})}{t_{PHSM,2}-t_{PHSM,1}}, & t_{PHSM,1} \le t < t_{PHSM,2} \\ \kappa_{a,1} + \frac{(\kappa_{a,2}-\kappa_{a,1})(t-t_{PHSM,2})}{t_{PHSM,3}-t_{PHSM,2}}, & t_{PHSM,2} \le t < t_{PHSM,3} \\ \kappa_{a,2} + \frac{(\kappa_{a,3}-\kappa_{a,2})(t-t_{PHSM,3})}{t_{PHSM,4}-t_{PHSM,3}}, & t_{PHSM,3} \le t < t_{PHSM,4} \\ \kappa_{a,3} + \frac{(\kappa_{a,4}-\kappa_{a,3})(t-t_{PHSM,4})}{t_{PHSM,5}-t_{PHSM,4}}, & t_{PHSM,4} \le t < t_{PHSM,5} \\ \kappa_{a,4} + \frac{(\kappa_{a,5}-\kappa_{a,4})(t-t_{PHSM,5})}{t_{PHSM,6}-t_{PHSM,5}}, & t_{PHSM,5} \le t < t_{PHSM,6} \end{cases}$$

Here:

- $t_{PHSM,1}$: Jan 23, 2020 — implementation of stringent PHSMs during the first COVID-19 wave in Hong Kong.
- $t_{PHSM,2}$: Mar 1, 2022 — PHSMs further strengthened during the fifth Omicron wave.
- $t_{PHSM,3}$: Sep 1, 2022 — major relaxation of community PHSMs and full-day school resumption.
- $t_{PHSM,4}$: Apr 1, 2023 — most PHSMs relaxed following the reopening of mainland China.
- $t_{PHSM,5}$: May 1, 2024 — local and international travel returned to pre-pandemic levels.

- $t_{PHSM,6}$: Apr 30, 2025 — end of simulation.

***Parameter estimation***

We performed parameter inference on the set $\theta_j$ (**Table S2**) for each region $j$: Beijing, Hong Kong, and Thailand. We assumed that the observed age-specific monthly hospitalizations or laboratory-confirmed cases $y^a_{j,m_k}$ follow a binomial distribution. Thus, the likelihood function of region $j$ is formulated as:

$$L_{case,j}(\theta_j) = \prod_{m_k} \prod_{a=1}^{25} Binomial(y^a_{j,m_k}; x^a_{j,m_k}, \epsilon_a)$$

In the case of Hong Kong, non-age-specific weekly laboratory-confirmed cases $y_{HK,w_k}$ were additionally available from January 2014, and the likelihood is:

$$L_{case,HK}(\theta_j) = \prod_{w_k} Poisson\left(y_{HK,w_k}; \sum_{a=1}^{25} \epsilon_a x^a_{HK,w_k}\right) \times \prod_{m_k} \prod_{a=1}^{25} Binomial(y^a_{HK,m_k}; x^a_{HK,m_k}, \epsilon_a)$$

As noted above, for each of the three regions, we calibrated the RSV transmission model to region-specific hospital admission or laboratory-confirmed case data while incorporating all available seroprevalence data. We assumed that, at equilibrium, the age-specific RSV seroprevalence $p^a_s$ remained constant over time $t$ during the period 2014–2019. For a given seroprevalence study $z$, we assumed that the observed number of seropositive samples $n^a_{z,positive}$ in age group $a$ among $n^a_{z,test}$ tested samples followed a binomial distribution. Accordingly, the likelihood function for all seroprevalence studies in region $j$ was:

$$L_{sero,j}(\theta_j) = \prod_{z} \prod_{a} Binomial(n^a_{z,positive}; n^a_{z,test}, p^a_s)$$

The overall likelihood function for each region $j$ is

$$L_j(\theta_j) = L_{case,j}(\theta_j) \times L_{sero,j}(\theta_j)$$

Using the likelihood and the prior distributions (**Table S2**), the posterior distribution of parameters in the model were determined by performing statistical inference in a Bayesian framework using Markov Chain Monte Carlo with Gibbs sampling [27].

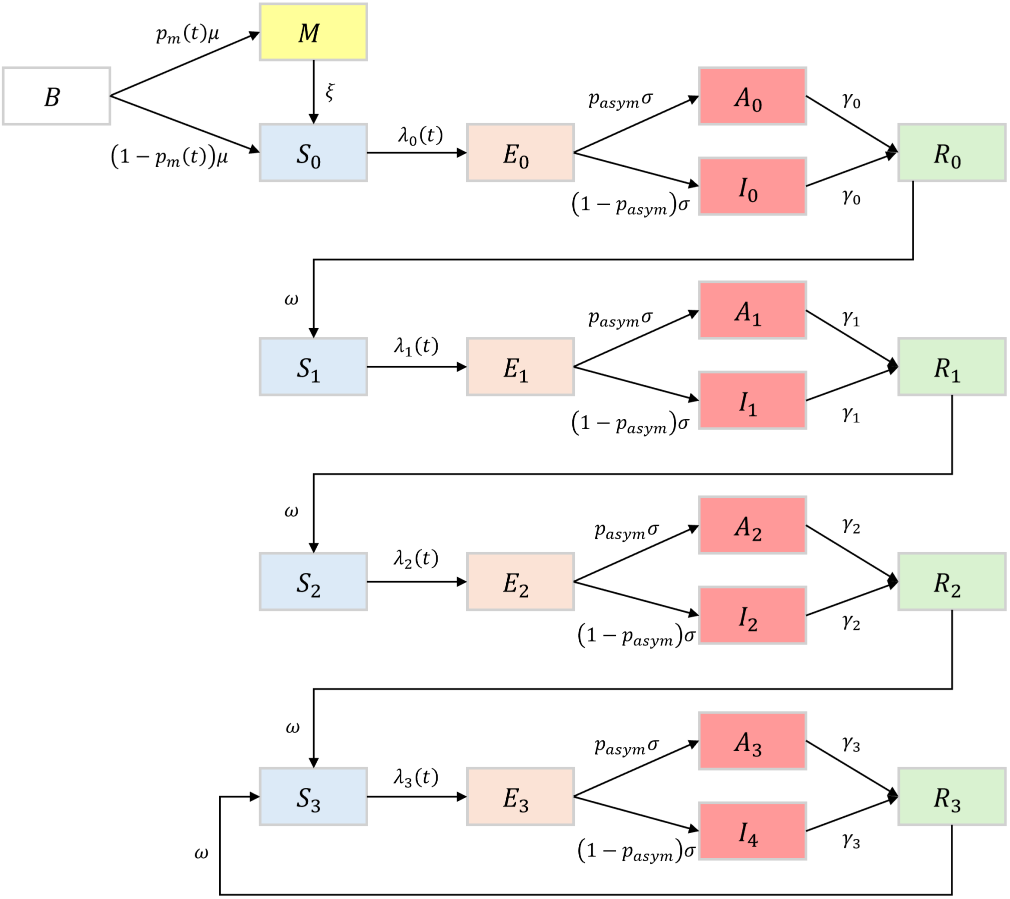


**Figure S1. Model diagram.** Epidemiological states are all age-specific ($B$: new births; $M$: completely protected due to maternally-derived immunity; $S$: susceptible; $E$: infected but not yet infectious; $A$: infectious and asymptomatic; $I$: infectious and symptomatic; $R$: recovered and protected due to infection) for each of the four levels stratified by the number of prior infections ($i \in \{0,1,2,3\}$). $\mu$ is the birth rate. $p_m(t)$ is the proportion of newborns who are completely protected by maternally derived immunity. $\xi$ is the waning rate of maternal protection. $\lambda_i(t)$ is the force of infection at time $t$ for level $i$. $\sigma$ is the rate of becoming infectious after infection. $p_{asym}$ is the age-specific proportion of asymptomatic infection. $\gamma_i$ is the rate of recovery for level $i$, and $\omega$ is the waning rate of the infection-induced protection.

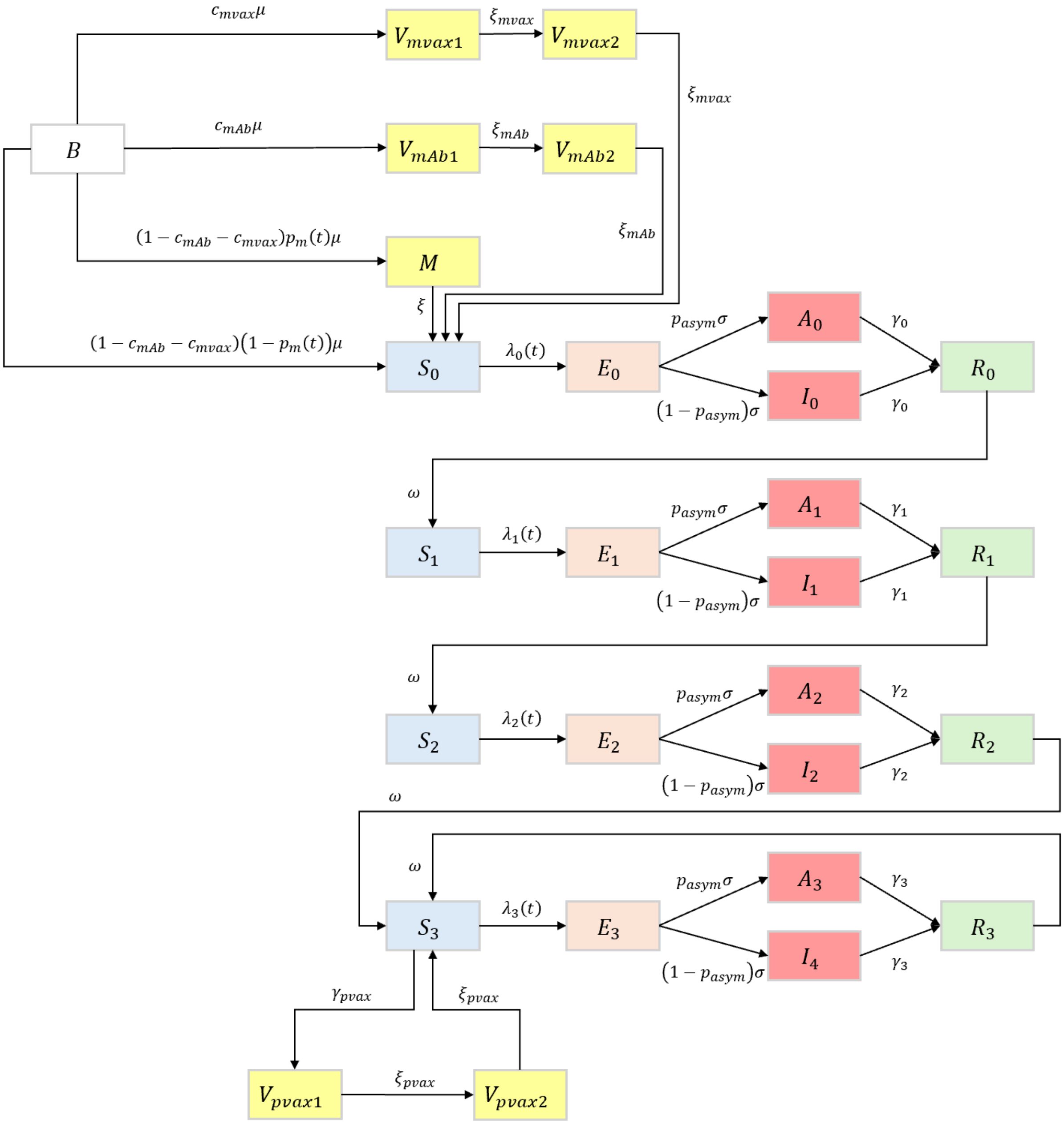


**Figure S2. Model diagram with interventions.** The model incorporates three interventions: (i) administration of long-acting monoclonal antibodies (mAbs), represented by compartments $V_{mAb1}$ and $V_{mAb2}$; (ii) maternal vaccination, represented by compartments $V_{mvax1}$ and $V_{mvax2}$; and (iii) vaccination of older adults, represented by compartments $V_{pvax1}$ and $V_{pvax2}$. Coverage for mAbs and maternal vaccination is denoted by $c_{mAb}$ and $c_{mvax}$, respectively, while $\gamma_{pvax}$ denotes the vaccination rate for older adults. The mean durations of protection are estimated from the published vaccine effectiveness, which are denoted with $^2/_{\xi_{mAb}}$, $^2/_{\xi_{mvax}}$, and $^2/_{\xi_{pvax}}$, respectively.

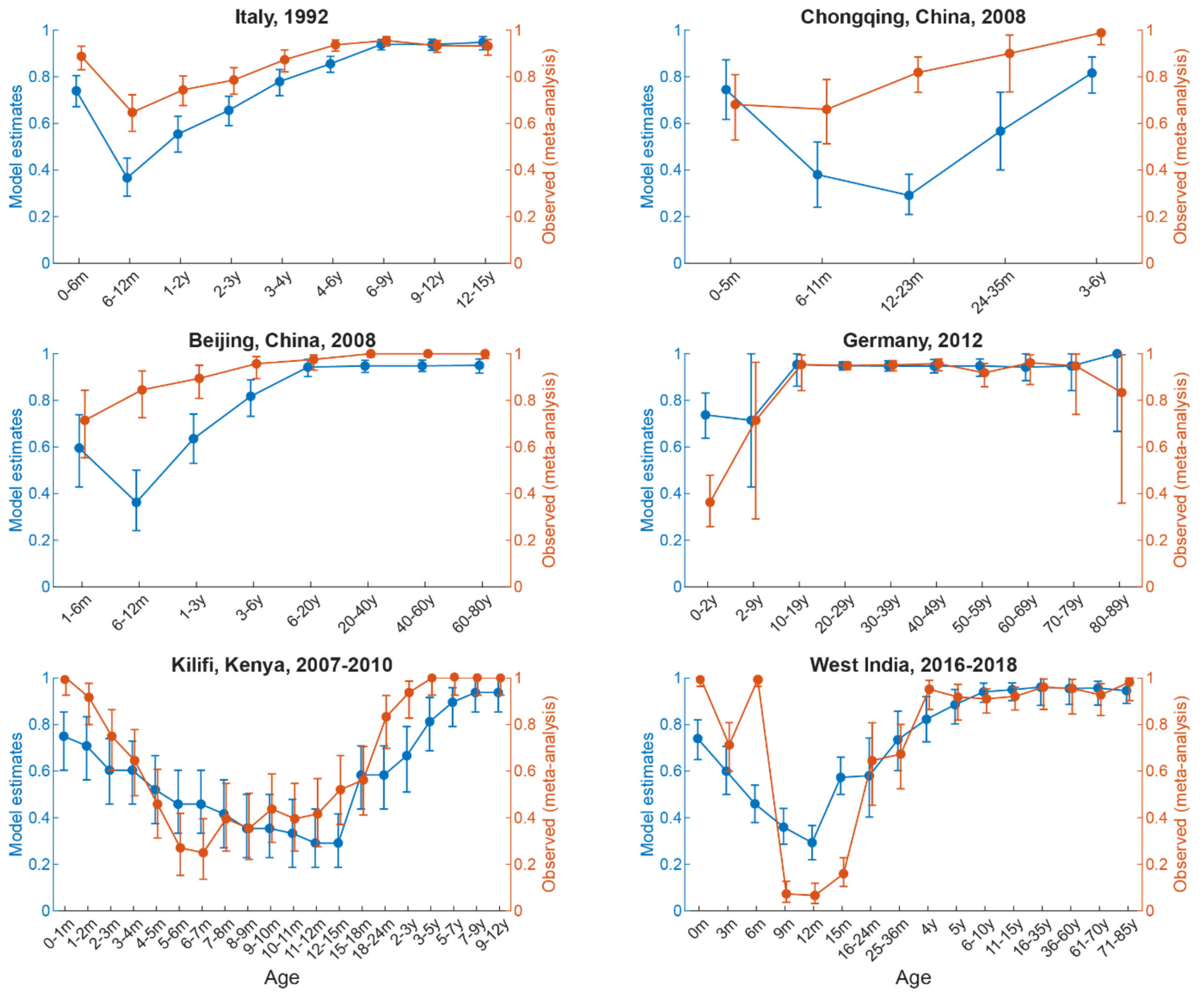


**Figure S3. Estimated RSV seroprevalence by age in Hong Kong between 2014 and 2019 and model fit to seroprevalence data summarised in the meta-analysis by Nakajo and Nishiura.** The age-specific seroprevalences from the Hong Kong transmission model were fit to all available seroprevalence data [17-19]. Blue triangles indicated the posterior mean, and blue bars indicated 95% CrI. Orange dots indicated the observed seroprevalence, and orange bars indicated the 95% CI assuming the number of seropositive samples follow binomial distribution.

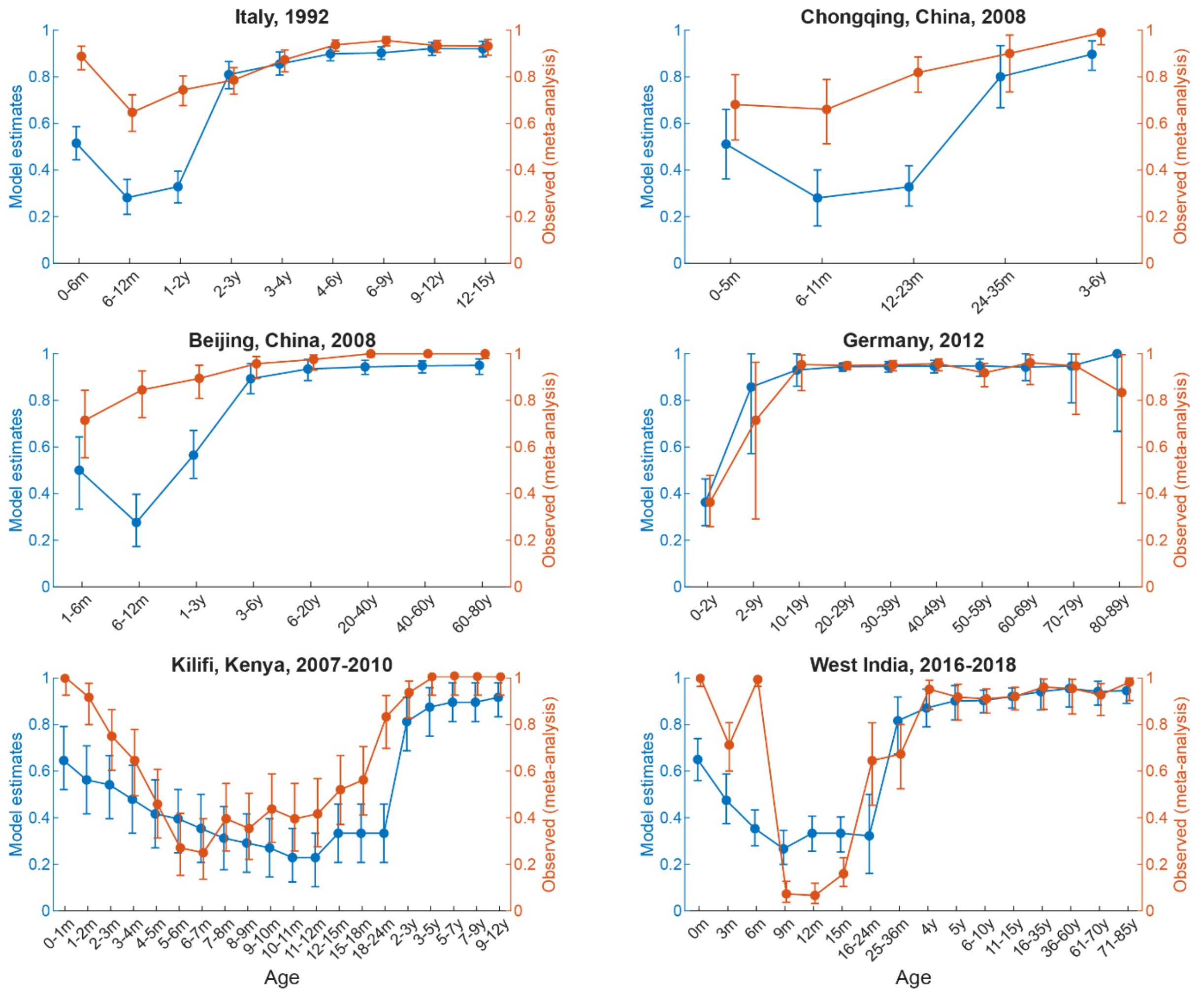


**Figure S4. Estimated RSV seroprevalence by age in Beijing between 2014 and 2019 and model fit to seroprevalence data summarised in the meta-analysis by Nakajo and Nishiura.** The age-specific seroprevalences from the Beijing transmission model were fit to all available seroprevalence data [17-19]. Blue triangles indicated the posterior mean, and blue bars indicated 95% CrI. Orange dots indicated the observed seroprevalence, and orange bars indicated the 95% CI assuming the number of seropositive samples follow binomial distribution.

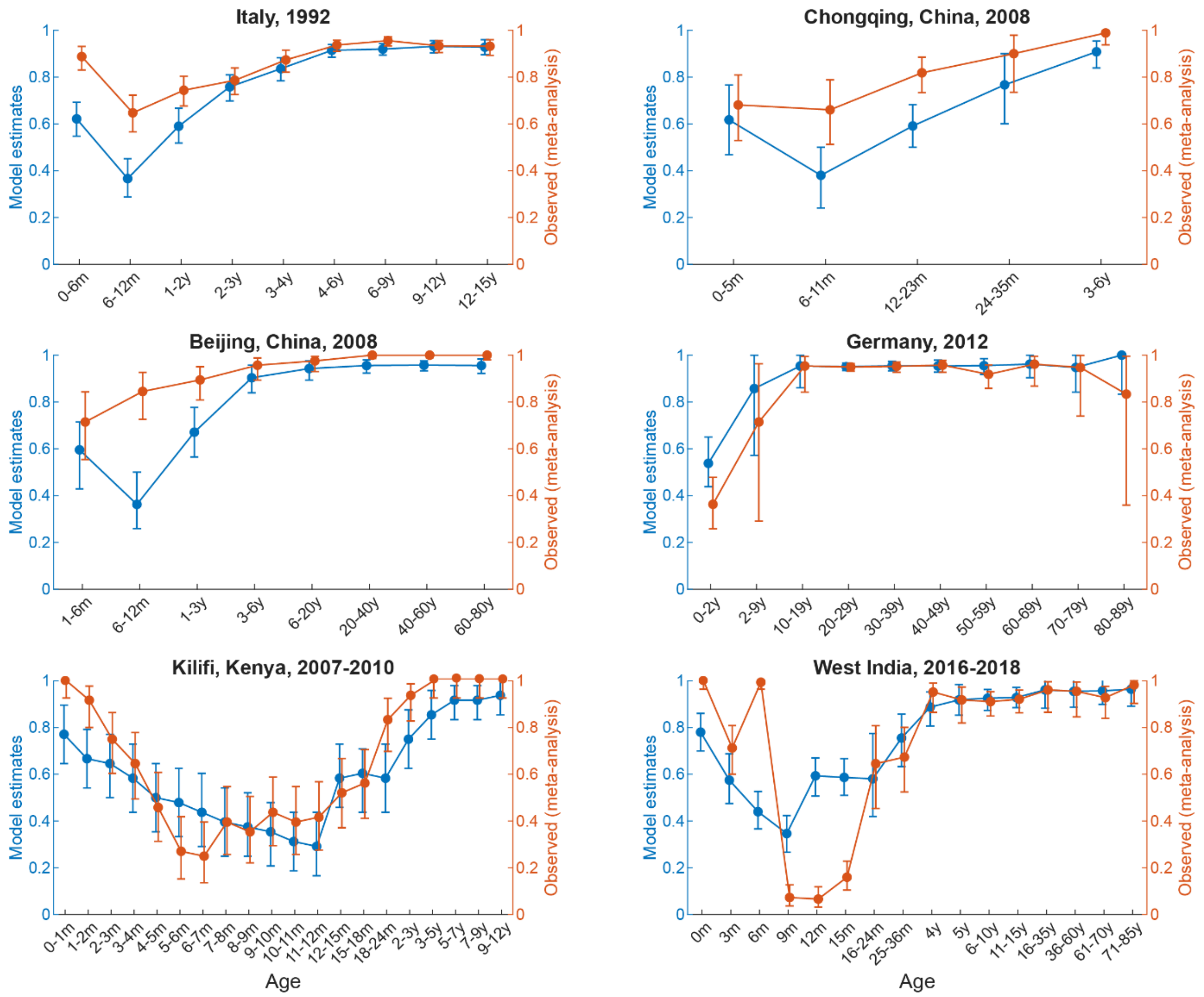


**Figure S5. Estimated RSV seroprevalence by age in Thailand between 2014 and 2019 and model fit to seroprevalence data summarised in the meta-analysis by Nakajo and Nishiura.** The age-specific seroprevalences from the Thailand transmission model were fit to all available seroprevalence data [17-19]. Blue triangles indicated the posterior mean, and blue bars indicated 95% CrI. Orange dots indicated the observed seroprevalence, and orange bars indicated the 95% CI assuming the number of seropositive samples follow binomial distribution.

Monthly RSV cases per 100,000 by age groups

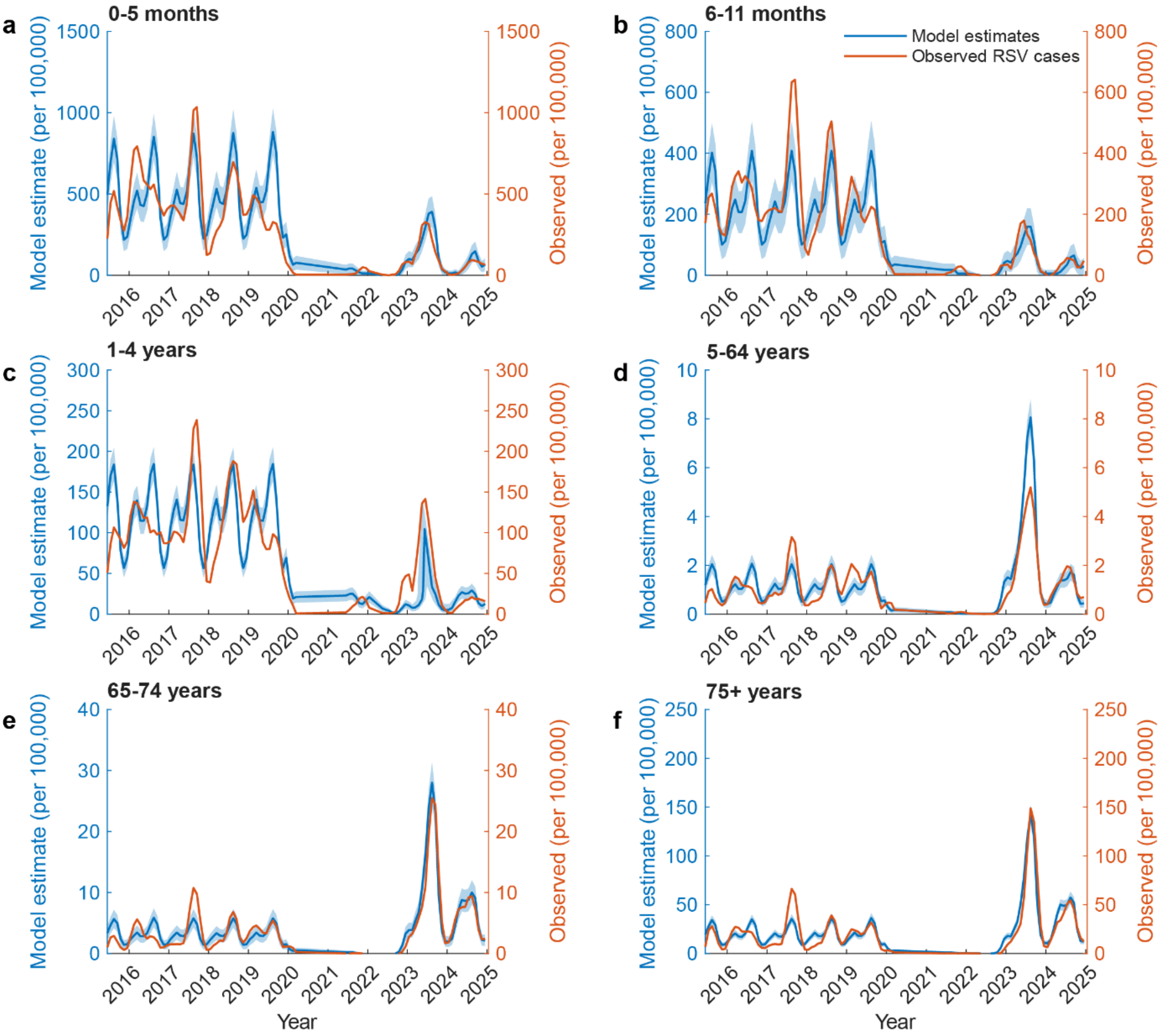


**Figure S6. Monthly number of laboratory-confirmed RSV cases per 100,000 population by age in Hong Kong between 1 June 2015 to 31 December 2024.** (a) 0-5 months. (b) 6-11 months. (c) 1-4 years. (d) 5-64 years. (e) 65-74 years. (f) ≥ 75 years. Blue solid lines indicated posterior mean, and blue shades indicated 95% CrI. Orange lines indicated the observed monthly number of laboratory-confirmed RSV cases per 100,000 population (3-month moving average accounting for reporting delays [28]). The x-axis labels indicated January 1 of each year.

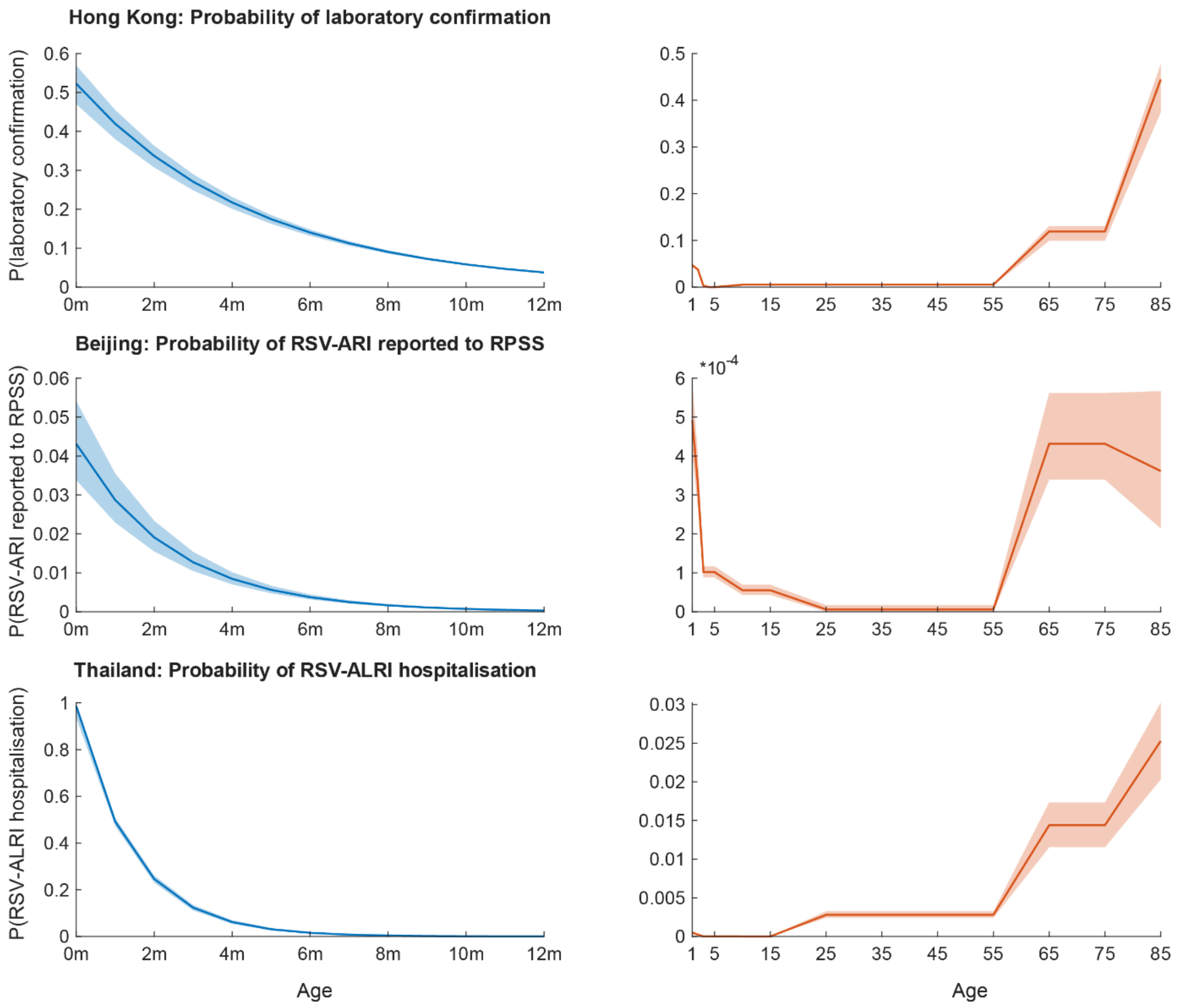


**Figure S7. Probability of RSV infection being reported and tested by age between 2014 and 2019 in Hong Kong, Beijing, and Thailand.** For Hong Kong, the probability of laboratory-confirmed RSV infection was estimated; for Beijing, the probability of RSV-associated ARI being reported to RPSS; and for Thailand, the probability of RSV-associated ALRI hospitalization. Blue solid lines represent posterior means for infants aged ≤1 year, with blue shading indicating the 95% CrI. Orange solid lines represent posterior means for individuals aged >1 year, with orange shading indicating the 95% CrI.

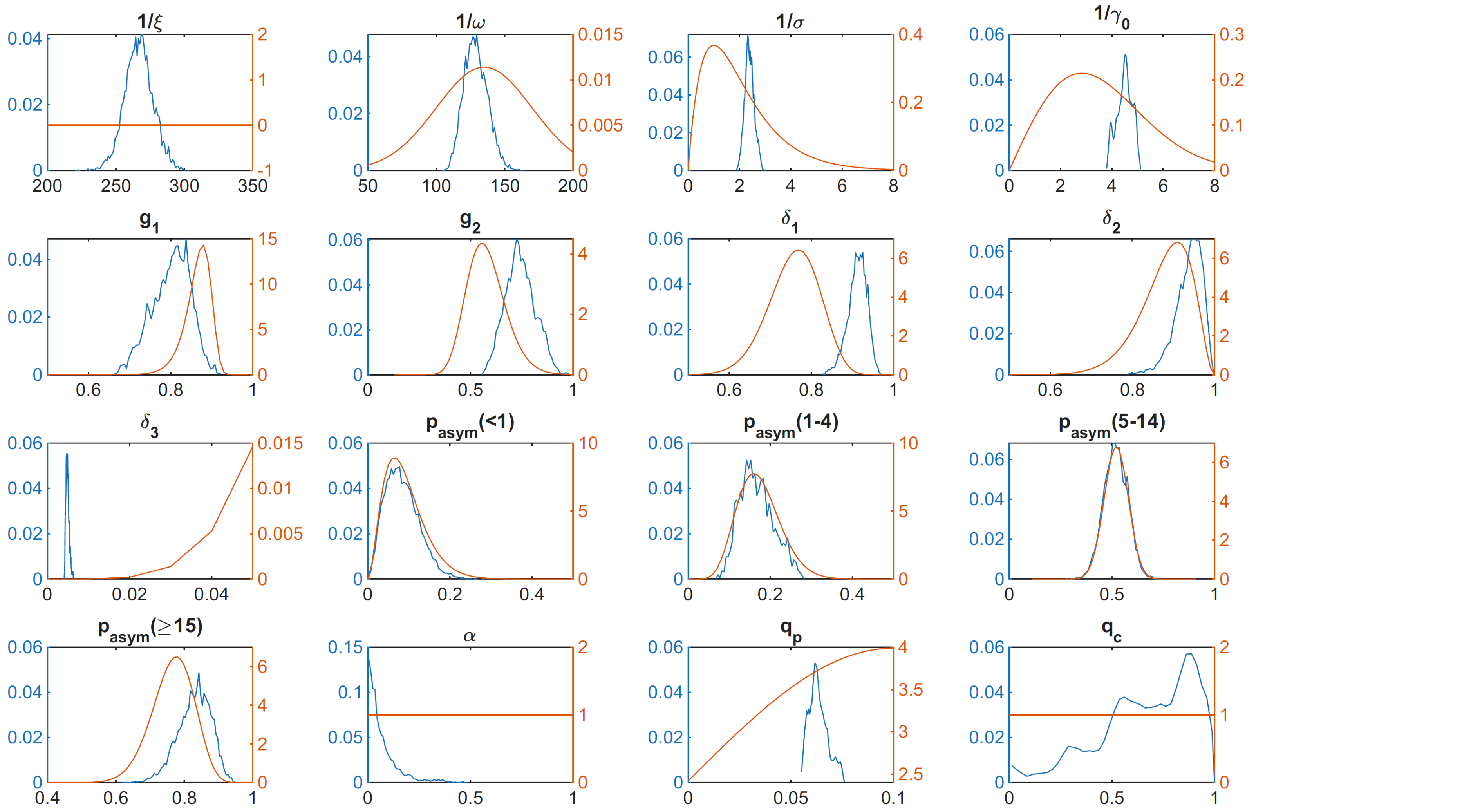


**Figure S8. Posterior distributions of parameters from the Hong Kong transmission model.** Empirical probability density functions of the posterior distributions are shown in blue for key parameters inferred during model calibration, while the corresponding prior distributions are shown in orange.

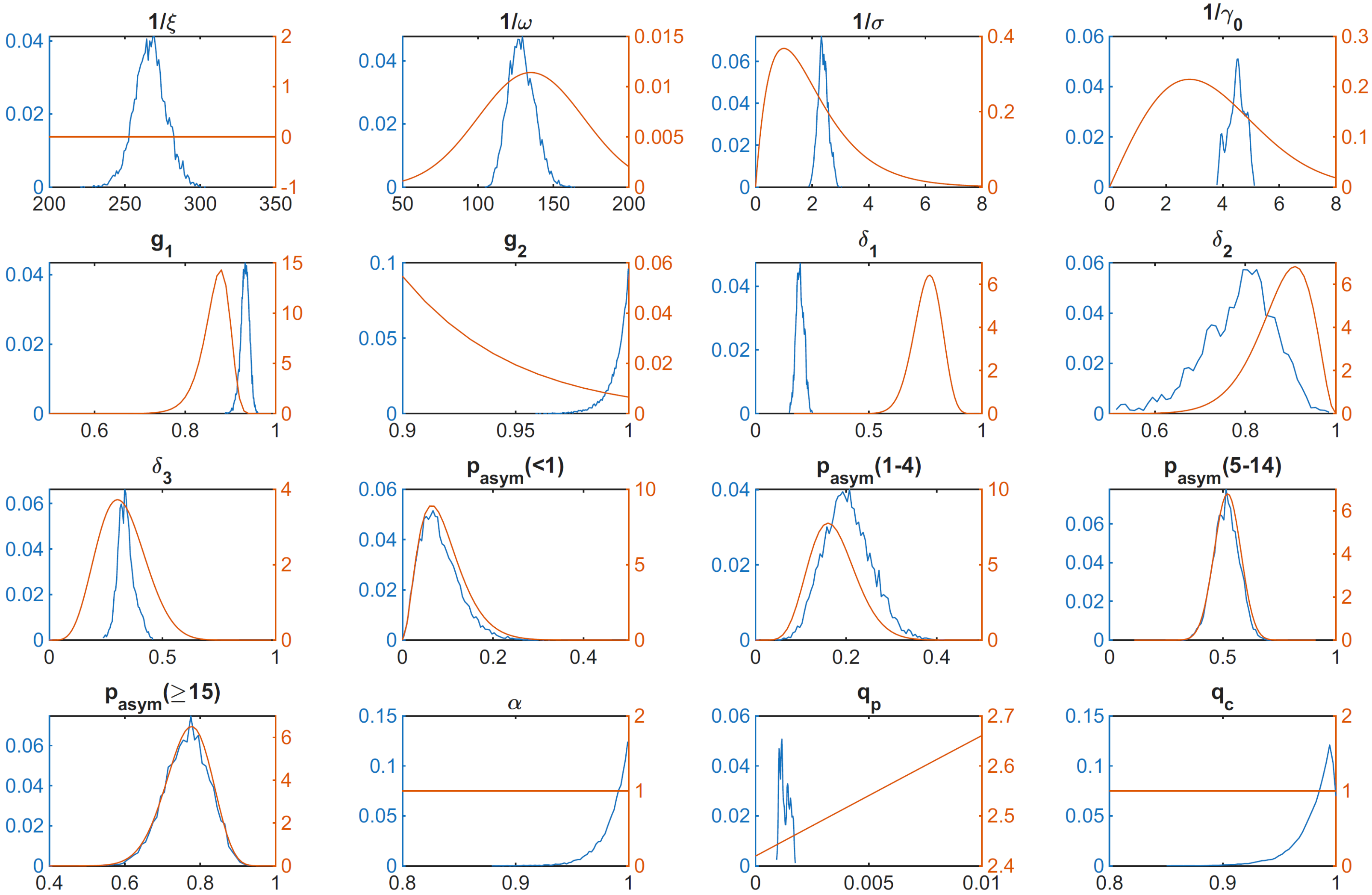


**Figure S9. Posterior distributions of parameters from the Beijing transmission model.** Empirical probability density functions of the posterior distributions are shown in blue for key parameters inferred during model calibration, while the corresponding prior distributions are shown in orange.

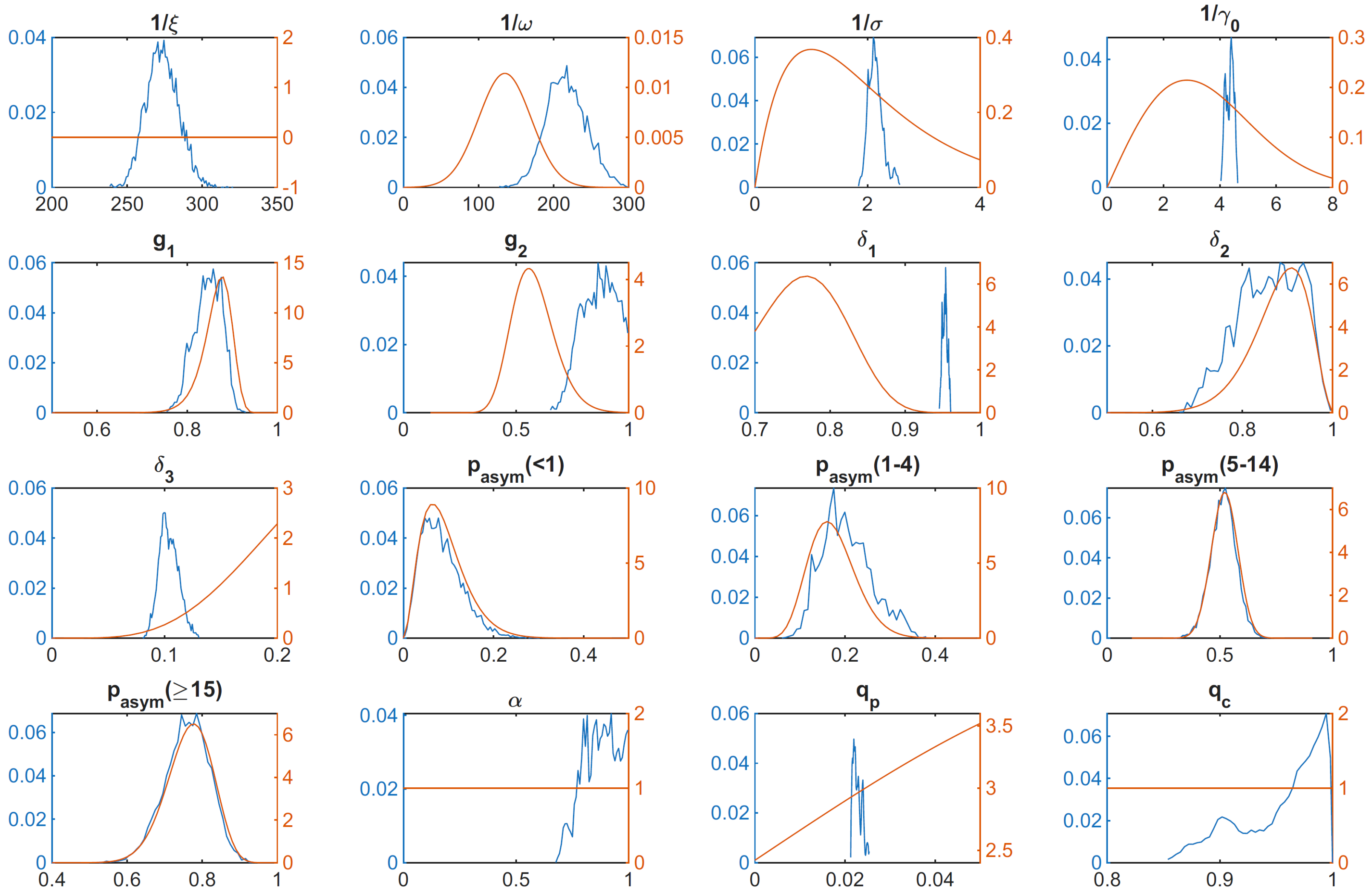


**Figure S10 Posterior distributions of parameters from the Thailand transmission model.** Empirical probability density functions of the posterior distributions are shown in blue for key parameters inferred during model calibration, while the corresponding prior distributions are shown in orange.

**Table S1. Epidemiological state variables of the RSV transmission model**

| State* | Description |
|---|---|
| $M^a(t)$ | Number of individuals aged $a$ at time $t$ who are completely protected from infection by maternally derived immunity |
| $S_i^a(t)$ | Number of individuals aged $a$ at time $t$ who are susceptible to infection, who have experienced $i$ infections previously |
| $E_i^a(t)$ | Number of individuals aged $a$ at time $t$ who are infected by RSV but have not yet been infectious, who have experienced $i$ infections previously |
| $A_i^a(t)$ | Number of individuals aged $a$ at time $t$ who are infected by RSV, infectious but have no symptoms, who have experienced $i$ infections previously |
| $I_i^a(t)$ | Number of individuals aged $a$ at time $t$ who are infected by RSV, infectious and have symptoms, who have experienced $i$ infections previously |
| $R_i^a(t)$ | Number of individuals aged $a$ at time $t$ who are completely protected from infection for a short time due to sterilising immunity acquired from natural infection, who have experienced $i$ infections previously |
| $V_{mvax1}^a(t)$<br>$V_{mvax2}^a(t)$ | Number of individuals aged $a$ at time $t$ who are completely protected from infection by maternal vaccination. We split the immunised state into two successive compartment and assumed that the mean protection duration follows Erlang distribution. |
| $V_{mAb1}^a(t)$<br>$V_{mAb2}^a(t)$ | Number of individuals aged $a$ at time $t$ who are completely protected from infection by administration of mAbs. We split the immunised state into two successive compartment and assumed that the mean protection duration follows Erlang distribution. |
| $V_{pvax1}^a(t)$<br>$V_{pvax2}^a(t)$ | Number of individuals aged $a$ at time $t$ who are partially protected from infection by vaccinating older adults. We split the immunised state into two successive compartment and assumed that the mean protection duration follows Erlang distribution. |
| $X^a(t)$ | Cumulative number of RSV infections from age group $a$ at time $t$ |

*$a \in \{1,2,\ldots,25\}$ and $i \in \{0,1,2,3\}$

**Table S2. Fixed parameters of the transmission models**

| Parameter | Description | Source |
|---|---|---|
| $\mu$ | Daily number of new births | Fixed [2-4] |
| $\eta$ | Aging rate from age group $a$ to $a+1$ | Fixed [2-4] |
| $c_{phy}^{a,b}$ | Daily rate of physical contacts made by age group $a$ with age group $b$ | Fixed [5-7] |
| $c_{con}^{a,b}$ | Daily rate of conversational contacts made by age group $a$ with age group $b$ | Fixed [5-7] |
| $c_{mAb}^{a}$ | Age-specific coverage of long-acting mAbs (i.e., 25% of RSV-associated severe outcomes attributed to all preterm births eligible for mAbs) | 0.25 [12] |
| $c_{mvax}^{a}$ | Age-specific coverage of maternal vaccination | 0.385 [15] |
| $\gamma_{pvax}^{a}$ | Age-specific rate of vaccination for older adults when they get one year older (to achieve uptake of 0.314 and 0.397 within 180 days for 60-74 and ≥75 years old) | 0.314 for 60-74 years<br>0.397 for ≥75 years |
| $p_{mAb}$ | Effectiveness of the long-acting mAbs within 180 days | 0.773 (0.503-0.897) [13] |
| $p_{mvax}$ | Effectiveness of maternal vaccination within 180 days | 0.694 (0.443-0.841) [16] |
| $p_{pvax}$ | Effectiveness of vaccinating older adults within 180 days | 0.80 (0.71-0.85) [22] |
| $2/\xi_{mAb}$ | Mean duration of protection of the long-acting mAbs | Estimated from $p_{mAb}$ |
| $2/\xi_{mvax}$ | Mean duration of protection of maternal vaccination | Estimated from $p_{mvax}$ |
| $2/\xi_{pvax}$ | Mean duration of protection of vaccinating older adults | Estimated from $p_{pvax}$ |

**Table S3. Prior distributions for parameters estimated in the inference**

| **Parameters estimated for Hong Kong, Beijing and Thailand** | | |
|---|---|---|
| **Parameter** | **Description** | **Prior distribution** |
| $1/\xi$ | Duration of maternally derived immunity (days) | $U$(14, 1825) |
| $I_1(0)$ | Initial proportion of people who are infected at $t = 0$ on 1 May 2004 (in compartments $E$, $A$, $I$) | $U$(0, 1) |
| $I_2(0)$ | Initial proportion of people who have recovered at $t = 0$ on 1 May 2004 (in compartment $R$) | $U$(0, 1) |
| $1/\omega$ | Mean duration of post-infection immunity (days) | $N$(365, 35) |
| $1/\sigma$ | Mean duration of latent period (days) | $Gamma$(7.1, 0.6) [29] |
| $1/\gamma_0$ | Mean duration of primary infection (days) | $Weibull$(4.1, 8.3) [30] |
| $g_1$ | Mean duration of secondary infection relative to primary infection | $Weibull$(34.2, 0.9) [30] |
| $g_2$ | Mean duration of subsequent infection relative to secondary infection | $LogN$(-0.56, 0.16) [1] |
| $\delta_0$ | Susceptibility to primary infection | 1 |
| $\delta_1$ | Susceptibility to secondary infection (relative to primary infection) | $Beta$(35.6, 11.4) [31] |
| $\delta_2$ | Susceptibility to tertiary infection (relative to secondary infection) | $Beta$(22.8, 3.2) [31] |
| $\delta_3$ | Susceptibility to subsequent infection (relative to tertiary infection) | $Beta$(6.1, 12.9) [31] |
| $p_{asym}^{<1}$ | Proportion of asymptomatic infections (<1 yr) | $U$(0, 1) |
| $p_{asym}^{1-4}$ | Proportion of asymptomatic infections (1-4 yr) | $U$(0, 1) |
| $p_{asym}^{5-14}$ | Proportion of asymptomatic infections (5-14 yr) | $U$(0, 1) |
| $p_{asym}^{\geq 15}$ | Proportion of asymptomatic infections (≥15 yr) | $U$(0, 1) |
| $\alpha$ | Relative infectiousness for asymptomatic infections | $U$(0, 1) |
| $q_p$ | Probability of RSV transmission per physical contact | $U$(0, 1) |
| $q_c$ | Relative RSV transmission per conversational contact | $U$(0, 1) |
| $b_1$ | Relative amplitude of transmission compared to peak | $U$(0, 5) |
| $\varphi$ | Seasonal shift in transmission | $U$(0, 1) |
| $\psi$ | Seasonality wavelength constant | $U$(0, 1) |
| $\epsilon_\lambda$ | Probability that an RSV infection is reported by CHP (0-4 yr), expressed by $e^{\epsilon_\lambda x_a + \epsilon_c}$ where $x_a$ is the age of years | $U$(-10, 0) |
| $\epsilon_c$ | | $U$(-10, 0) |
| $\epsilon_{5-54}$ | Probability that an RSV infection is reported and tested (5-54 yr) | $U$(0, 1) |
| $\epsilon_{55-74}$ | Probability that an RSV infection is reported and tested (55-64 yr) | $U$(0, 1) |
| $\epsilon_{\geq 75}$ | Probability that an RSV infection is reported and tested (≥ 75 yr) | $U$(0, 1) |

| | | |
|---|---|---|
| $\pi$ | Probability of IgG ELISA seropositivity immediately after RSV infection | $U(0, 1)$ |
| **Parameters estimated for Hong Kong only with data from 2020 to 2024** | | |
| **Parameter** | **Description** | **Prior distribution** |
| $\kappa_{a,1}$ | Relative reduction or increase in RSV transmissibility on 2022-03-01, compared with 2014-2019, $a \in \{< 1, 1-4, 5-64, 65-74, \geq 75\}$ | $U(0, 5)$ |
| $\kappa_{a,2}$ | Relative reduction or increase in RSV transmissibility on 2022-09-01, compared with 2014-2019, $a \in \{< 1, 1-4, 5-64, 65-74, \geq 75\}$ | $U(0, 5)$ |
| $\kappa_{a,3}$ | Relative reduction or increase in RSV transmissibility on 2023-04-01, compared with 2014-2019, $a \in \{< 1, 1-4, 5-64, 65-74, \geq 75\}$ | $U(0, 5)$ |
| $\kappa_{a,4}$ | Relative reduction or increase in RSV transmissibility on 2024-05-01, compared with 2014-2019, $a \in \{< 1, 1-4, 5-64, 65-74, \geq 75\}$ | $U(0, 5)$ |
| $\kappa_{a,5}$ | Relative reduction or increase in RSV transmissibility on 2025-04-30, compared with 2014-2019, $a \in \{< 1, 1-4, 5-64, 65-74, \geq 75\}$ | $U(0, 5)$ |

**Table S4. Posterior distribution of model parameters estimated in the inference**

| Parameter | Description | Hong Kong (95% CrI) | Beijing (95% CrI) | Thailand (95% CrI) |
|---|---|---|---|---|
| $1/\xi$ | Mean duration of maternally derived immunity (days) | 267 (246 – 288) | 269 (243 – 286) | 273 (253 – 295) |
| $1/\omega$ | Mean duration of post-infection immunity (days) | 128 (113 – 147) | 117 (101 – 136) | 216 (136 – 271) |
| $1/\sigma$ | Mean duration of exposure (days) | 2.38 (2.05 – 2.77) | 2.48 (2.14 – 2.68) | 2.13 (1.93 – 2.48) |
| $1/\gamma_0$ | Mean duration of primary infection (days) | 4.51 (3.91 – 4.99) | 4.43 (3.82 – 4.91) | 4.37 (4.10 – 4.58) |
| $g_1$ | Duration of secondary infection relative to primary infection | 0.81 (0.70 – 0.88) | 0.93 (0.91 – 0.95) | 0.85 (0.79 – 0.90) |
| $g_2$ | Duration of subsequent infection relative to secondary infection | 0.74 (0.61 – 0.89) | 0.99 (0.98 – 1.00) | 0.87 (0.72 – 0.99) |
| $\delta_1$ | Susceptibility to secondary infection (relative to primary infection) | 0.91 (0.86 – 0.95) | 0.20 (0.16 – 0.23) | 0.047 (0.041 – 0.053) |
| $\delta_2$ | Susceptibility to tertiary infection (relative to secondary infection) | 0.94 (0.86 – 0.99) | 0.80 (0.60 – 0.92) | 0.86 (0.72 – 0.97) |
| $\delta_3$ | Susceptibility to subsequent infection (relative to tertiary infection) | 0.05 (0.04 – 0.06) | 0.34 (0.27 – 0.42) | 0.10 (0.09 – 0.12) |
| $p_{asym}^{<1}$ | Proportion of asymptomatic infections (<1 yr) | 0.079 (0.020 – 0.172) | 0.075 (0.018 – 0.186) | 0.078 (0.019 – 0.188) |
| $p_{asym}^{1-4}$ | Proportion of asymptomatic infections (1-4 yr) | 0.162 (0.097 – 0.254) | 0.200 (0.110 – 0.314) | 0.108 (0.113 – 0.327) |
| $p_{asym}^{5-14}$ | Proportion of asymptomatic infections (5-14 yr) | 0.518 (0.404 – 0.629) | 0.516 (0.406 – 0.622) | 0.517 (0.404 – 0.626) |
| $p_{asym}^{\geq 15}$ | Proportion of asymptomatic infections (≥15 yr) | 0.833 (0.725 – 0.910) | 0.767 (0.642 – 0.870) | 0.763 (0.638 – 0.867) |
| $\alpha$ | Relative infectiousness for asymptomatic infections | 0.955 (0.702 – 0.998) | 0.990 (0.948 – 1.000) | 0.876 (0.718 – 0.994) |
| $q_p$ | Probability of RSV transmission per physical contact | 0.063 (0.056 – 0.074) | 0.013 (0.001 – 0.07) | 0.023 (0.021 – 0.025) |
| $q_c$ | Relative RSV transmission per conversational contact | 0.708 (0.140 – 0.983) | 0.987 (0.949 – 1.000) | 0.968 (0.872 – 0.999) |
| $b_1$ | Relative amplitude of transmission during peak | 0.847 (0.612 – 1.161) | 9.50 (6.66 – 11.86) | 1.17 (1.06 – 1.25) |

| | | | | |
|---|---|---|---|---|
| $\varphi$ | Seasonal shift in transmission | 0.533 (0.529 – 0.537) | 0.994 (0.971 – 1.000) | 0.33 (0.32 – 0.34) |
| $\psi$ | Seasonality wavelength constant | 0.458 (0.426 – 0.495) | 0.988 (0.949 – 1.000) | 0.99 (0.96 – 1.00) |
| $\epsilon_\lambda$<br>$\epsilon_c$ | Probability that an RSV infection is reported by CHP (0-2 yr), expressed by $e^{\epsilon_\lambda x_a + \epsilon_c}$ where $x_a$ is the age of years | $3.03 \times \exp(-1.74 - 2.63x_a)$ | $\exp(-3.14 - 4.89x_a)$ | $\exp(-0.014 - 8.30x_a)$ |
| $\epsilon_{2-4}$ | Probability that an RSV infection is reported and tested (2-4 yr) * | | $101.8\ (88.3 - 116.8) \times 10^{-6}$ | $12.0\ (0.42 - 63.3) \times 10^{-6}$ |
| $\epsilon_{5-14}$ | Probability that an RSV infection is reported and tested (5-14 yr) * | 0.0053 (0.0041 – 0.0064) | $55.3\ (43.6 - 69.4) \times 10^{-6}$ | $5.45\ (0.18 - 26.8) \times 10^{-6}$ |
| $\epsilon_{15-54}$ | Probability that an RSV infection is reported and tested (5-54 yr) * | | $6.12\ (0.43 - 16.9) \times 10^{-6}$ | 0.0028 (0.0024 – 0.0032) |
| $\epsilon_{55-74}$ | Probability that an RSV infection is reported and tested (55-64 yr) * | 0.119 (0.0995 – 0.131) | 0.00043 (0.00034 – 0.00056) | 0.014 (0.012 – 0.017) |
| $\epsilon_{\geq 75}$ | Probability that an RSV infection is reported and tested (≥ 75 yr) * | 0.932 (0.786 – 0.997) | 0.00026 (0.000027 – 0.00057) | 0.042 (0.033 – 0.050) |
| $\pi$ | Probability of IgG ELISA seropositivity immediately after RSV infection | 0.952 (0.945 – 0.958) | 1.000 (0.990 – 1.000) | 0.960 (0.953 – 0.966) |

*Hong Kong: lab-confirmed RSV infections; Beijing: RSV-associated ARI reported to RPSS; Thailand: RSV-associated ALRI hospitalisations.

**Table S4. Estimated impacts of public health and social measures (PHSMs) on RSV transmission in Hong Kong, 2020–2024**

| Parameter | Description | Posterior mean (95% CrI) |
|---|---|---|
| $\kappa_{a,0}$ | Relative reduction or increase in RSV transmissibility on 2020-01-23, compared with 2014-2019. | 0-11 months: 1<br>1-4 years: 1<br>5-64 years: 1<br>65-74 years: 1<br>≥ 75 years: 1 |
| $\kappa_{a,1}$ | Relative reduction or increase in RSV transmissibility on 2022-03-01, compared with 2014-2019. | 0-11 months: 0.020 (0.011-0.032)<br>1-4 years: 0.195 (0.183-0.207)<br>5-64 years: 0.012 (0.002-0.025)<br>65-74 years: 0.003 (0.000-0.013)<br>≥ 75 years: 0.001 (0.000-0.004) |
| $\kappa_{a,2}$ | Relative reduction or increase in RSV transmissibility on 2022-09-01, compared with 2014-2019. | 0-11 months: 0.009 (0.000-0.034)<br>1-4 years: 0.054 (0.051-0.057)<br>5-64 years: 0.115 (0.089-0.142)<br>65-74 years: 0.009 (0.000-0.045)<br>≥ 75 years: 0.002 (0.000-0.012) |
| $\kappa_{a,3}$ | Relative reduction or increase in RSV transmissibility on 2023-04-01, compared with 2014-2019. | 0-11 months: 0.213 (0.192-0.237)<br>1-4 years: 0.029 (0.027-0.030)<br>5-64 years: 1.562 (1.509-1.617)<br>65-74 years: 1.428 (1.299-1.558)<br>≥ 75 years: 1.013 (0.947-1.078) |
| $\kappa_{a,4}$ | Relative reduction or increase in RSV transmissibility on 2024-05-01, compared with 2014-2019. | 0-11 months: 0.012 (0.004-0.022)<br>1-4 years: 0.076 (0.073-0.080)<br>5-64 years: 0.635 (0.596-0.673)<br>65-74 years: 1.370 (1.262-1.478)<br>≥ 75 years: 1.172 (1.115-1.228) |
| $\kappa_{a,5}$ | Relative reduction or increase in RSV transmissibility on 2025-04-30, compared with 2014-2019. | 0-11 months: 0.405 (0.324-0.491)<br>1-4 years: 0.196 (0.179-0.214)<br>5-64 years: 0.315 (0.162-0.471)<br>65-74 years: 0.078 (0.003-0.370)<br>≥ 75 years: 0.076 (0.004-0.282) |

### References


1. Hodgson D, Pebody R, Panovska-Griffiths J, Baguelin M, Atkins KE. Evaluating the next generation of RSV intervention strategies: a mathematical modelling study and cost-effectiveness analysis. BMC medicine. 2020;18:1–14.
2. Census and Statistics Department of The Government of the Hong Kong Special Administrative Region. Population and Household 2025. Available from: https://www.censtatd.gov.hk/en/.
3. National Bureau of Statistics of China. National data 2025. Available from: https://data.stats.gov.cn/.
4. Ministry of Public Health of Thailand. Public Health Statistics 2025. Available from: https://spd.moph.go.th/.
5. Leung K, Jit M, Lau EH, Wu JT. Social contact patterns relevant to the spread of respiratory infectious diseases in Hong Kong. Scientific reports. 2017;7(1):7974.
6. Mistry D, Litvinova M, Pastore y Piontti A, Chinazzi M, Fumanelli L, Gomes MF, et al. Inferring high-resolution human mixing patterns for disease modeling. Nature communications. 2021;12(1):323.
7. Prem K, Zandvoort Kv, Klepac P, Eggo RM, Davies NG, Group CftMMoIDC-W, et al. Projecting contact matrices in 177 geographical regions: an update and comparison with empirical data for the COVID-19 era. PLoS computational biology. 2021;17(7):e1009098.
8. Hansen CL, Lee L, Bents SJ, Perofsky AC, Sun K, Starita LM, et al. Scenario Projections of Respiratory Syncytial Virus Hospitalizations Averted Due to New Immunizations. JAMA network open. 2025;8(6):e2514622–e.
9. Pecenka C, Sparrow E, Feikin DR, Srikantiah P, Darko DM, Karikari-Boateng E, et al. Respiratory syncytial virus vaccination and immunoprophylaxis: realising the potential for protection of young children. The Lancet. 2024;404(10458):1157–70.
10. Hon K, Cheung EW, Li AM, Fung GP, Lam DS, Lee MS, et al. Practice recommendations for respiratory syncytial virus prophylaxis among children in Hong Kong. Hong Kong Medical Journal. 2025;31(1):48.
11. Deng K, Liang J, Mu Y, Liu Z, Wang Y, Li M, et al. Preterm births in China between 2012 and 2018: an observational study of more than 9 million women. The Lancet Global Health. 2021;9(9):e1226–e41.
12. Wang X, Li Y, Shi T, Bont LJ, Chu HY, Zar HJ, et al. Global disease burden of and risk factors for acute lower respiratory infections caused by respiratory syncytial virus in preterm infants and young children in 2019: a systematic review and meta-analysis of aggregated and individual participant data. The Lancet. 2024;403(10433):1241–53.

13. Hammitt LL, Dagan R, Yuan Y, Baca Cots M, Bosheva M, Madhi SA, et al. Nirsevimab for prevention of RSV in healthy late-preterm and term infants. New England Journal of Medicine. 2022;386(9):837–46.
14. Griffin MP, Yuan Y, Takas T, Domachowske JB, Madhi SA, Manzoni P, et al. Single-dose nirsevimab for prevention of RSV in preterm infants. New England Journal of Medicine. 2020;383(5):415–25.
15. Centers for Disease Control Prevention. Respiratory syncytial virus (RSV) vaccination coverage, pregnant persons, United States. 2024.
16. Kampmann B, Madhi SA, Munjal I, Simões EAF, Pahud BA, Llapur C, et al. Bivalent Prefusion F Vaccine in Pregnancy to Prevent RSV Illness in Infants. New England Journal of Medicine. 2023;388(16):1451–64. doi: doi:10.1056/NEJMoa2216480.
17. Kriss JL. Influenza, COVID-19, and Respiratory Syncytial Virus Vaccination Coverage Among Adults—United States, Fall 2024. MMWR Morbidity and Mortality Weekly Report. 2024;73.
18. Centre for Health Protection of The Government of the Hong Kong Special Administrative Region. Statistics on vaccination programmes in the past 3 years 2025. Available from: https://www.chp.gov.hk/en/features/102226.html.
19. Centers for Disease Control Prevention. Influenza Vaccination Coverage and Intent for Vaccination, Adults 18 Years and Older, United States 2025. Available from: https://www.cdc.gov/fluvaxview/dashboard/adult-coverage.html.
20. Shen Y, Wang J, Lv M, Wu J, Nicholas S, Maitland E, et al. Predicting future vaccination habits: The link between influenza vaccination patterns and future vaccination decisions among old aged adults in China. Journal of Infection and Public Health. 2024;17(6):1079–85.
21. Montgomery MP, Praphasiri P, Ditsungnoen D, Akarasewi P, Chittaganpitch M, Puthavathana P, et al. Influenza surveillance and vaccine policy in Thailand—a historical perspective. The Lancet Regional Health-Southeast Asia. 2025;41.
22. Payne AB, Watts JA, Mitchell PK, Dascomb K, Irving SA, Klein NP, et al. Respiratory syncytial virus (RSV) vaccine effectiveness against RSV-associated hospitalisations and emergency department encounters among adults aged 60 years and older in the USA, October, 2023, to March, 2024: a test-negative design analysis. The Lancet. 2024;404(10462):1547–59. doi: 10.1016/S0140-6736(24)01738-0.
23. Bloom-Feshbach K, Alonso WJ, Charu V, Tamerius J, Simonsen L, Miller MA, et al. Latitudinal variations in seasonal activity of influenza and respiratory syncytial virus (RSV): a global comparative review. PloS one. 2013;8(2):e54445.
24. Nakajo K, Nishiura H. Age-dependent risk of respiratory syncytial virus infection: A systematic review and hazard modeling from serological data. The Journal of Infectious Diseases. 2023;228(10):1400–9.

25. Lau YC, Ryu S, Du Z, Wang L, Wu P, Lau EH, et al. Impact of COVID-19 control measures on respiratory syncytial virus and hand-foot-and-mouth disease transmission in Hong Kong and South Korea. Epidemics. 2024;49:100797.

26. Yang B, Lin Y, Xiong W, Liu C, Gao H, Ho F, et al. Comparison of control and transmission of COVID-19 across epidemic waves in Hong Kong: an observational study. The Lancet Regional Health–Western Pacific. 2024;43.

27. Robert CP, Casella G, Casella G. Monte Carlo statistical methods: Springer; 1999.

28. Centre for Health Protection of The Government of the Hong Kong Special Administrative Region. Detection of pathogens from respiratory specimens 2025. Available from: https://www.chp.gov.hk/en/statistics/data/10/641/642/2274.html.

29. DeVincenzo JP, Wilkinson T, Vaishnaw A, Cehelsky J, Meyers R, Nochur S, et al. Viral load drives disease in humans experimentally infected with respiratory syncytial virus. American journal of respiratory and critical care medicine. 2010;182(10):1305–14.

30. Okiro EA, White LJ, Ngama M, Cane PA, Medley GF, Nokes DJ. Duration of shedding of respiratory syncytial virus in a community study of Kenyan children. BMC infectious diseases. 2010;10:1–7.

31. Henderson FW, Collier AM, Clyde Jr WA, Denny FW. Respiratory-syncytial-virus infections, reinfections and immunity: a prospective, longitudinal study in young children. New England Journal of Medicine. 1979;300(10):530–4.
